# Single-cell machine learning uncovers genetically anchored, cell-type specific programs of Alzheimer’s disease

**DOI:** 10.64898/2026.01.31.26345282

**Authors:** Adithya Madduri, Randall Ellis, Chirag Lakhani, David Bennett, Chirag J Patel

## Abstract

Aging and genetic risk shape the molecular programs that confer cellular vulnerability in Alzheimer’s disease (AD), but whether these programs differ between clinical symptoms and neuropathological burden remains unclear. Using single-nucleus RNA sequencing (snRNA-seq) from the dorsolateral prefrontal cortex of 48 donors either with AD or controls (∼70,000 nuclei), we integrated transcriptomic, genetic, and demographic data to identify shared and cell-type–specific molecular predictors of AD. Across six major brain cell types and 41 fine-grained subclusters, we uncovered robust pan-cell-type predictors—including *RASGEF1B*, *LINGO1*, and *ARL17B*—and distinct cell-specific programs such as *CRYAB* in oligodendrocytes and *IFI44L* in microglia. Many of these genes have regulatory links to genome-wide significant AD loci and brain eQTLs, linking genetic susceptibility to transcriptional state. Pseudotime analyses revealed progressive activation of amyloid and neuroinflammatory pathways along disease trajectories, while comparative modeling of clinical versus neuropathological outcomes highlighted divergent molecular programs between symptom manifestation and amyloid plaque burden. Validation in an independent cohort (21 donors, ∼172,000 nuclei) confirmed the reproducibility of predictive features across cell types. By jointly modeling genetic, demographic, and transcriptomic axes, our study nominates high-confidence, genetically anchored molecular drivers of AD and prioritizes them for mechanistic investigation and therapeutic targeting in age-related neurodegenerative disease.

## Introduction

Alzheimer’s disease (AD) is a progressive neurodegenerative disorder marked by the accumulation of amyloid-β plaques, intracellular neurofibrillary tangles (NFTs) in neurons, and cognitive decline, and is inextricably linked with age^1^ ^2^ ^3^ ^4^. Despite decades of research, AD continues to affect millions worldwide and remains a major public health challenge ^5,6^. Single-nucleus RNA sequencing (snRNA-seq) has provided foundational insights into the molecular pathology of AD, revealing cell-type-specific transcriptional alterations in neurons, astrocytes, and microglia ^7,8^. However, most studies to date have relied on univariate differential expression (DE) analyses, which fail to capture multivariate gene interactions that may underlie cellular vulnerability and disease progression.

At the same time, demographic and genetic risk factors such as age, sex, and APOE genotype are well known influencers of AD onset and severity ^9^ ^10^ ^11^. Yet how these variables interact with cell-type-specific molecular signatures remains poorly understood. Bridging this gap will require a more granular, cell-type–specific view of both clinical and neuropathological AD to inform the development of targeted diagnostics, prognostic tools, and therapeutic strategies.

To address this gap, we implemented a three-pronged analytical workflow, TriSCOPE, “Three-factor RNA-based System for Comprehensive Omics Profiling and Exploration”, an integrative machine learning framework that combines: (i) multivariate predictive modeling using AutoML, (ii) differential expression analysis, and (iii) Pseudotime-based trajectory inference. By incorporating orthogonal evidence beyond univariate fold change, TriSCOPE enables convergent validation of disease-relevant genes and reduces false positives ^12^. Although designed for broad application across single-cell datasets and multiple axes of disease, here we applied TriSCOPE to two Alzheimer’s cohorts. Applying TriSCOPE to 243,148 single-nucleus transcriptomes (Table 1) ^7,13,14^ from Dorsolateral Prefrontal Cortex (PFC)), a prefrontal region implicated in higher-order cognition and executive function ^15^, across two human brain cohorts, we identified robust, cell-type-specific predictors of both clinical AD diagnosis and neuropathological diagnosis of AD. We hypothesized that there may be divergent transcriptional programs between clinical and pathological manifestations of Alzheimer’s disease across brain cell types ^16,17^. Though clinical and pathological definitions are generally concordant in cohort settings, such as ROSMAP ^18^, underlying programs between the two definitions may diverge (Supplemental Table 16).

**Table 1:**
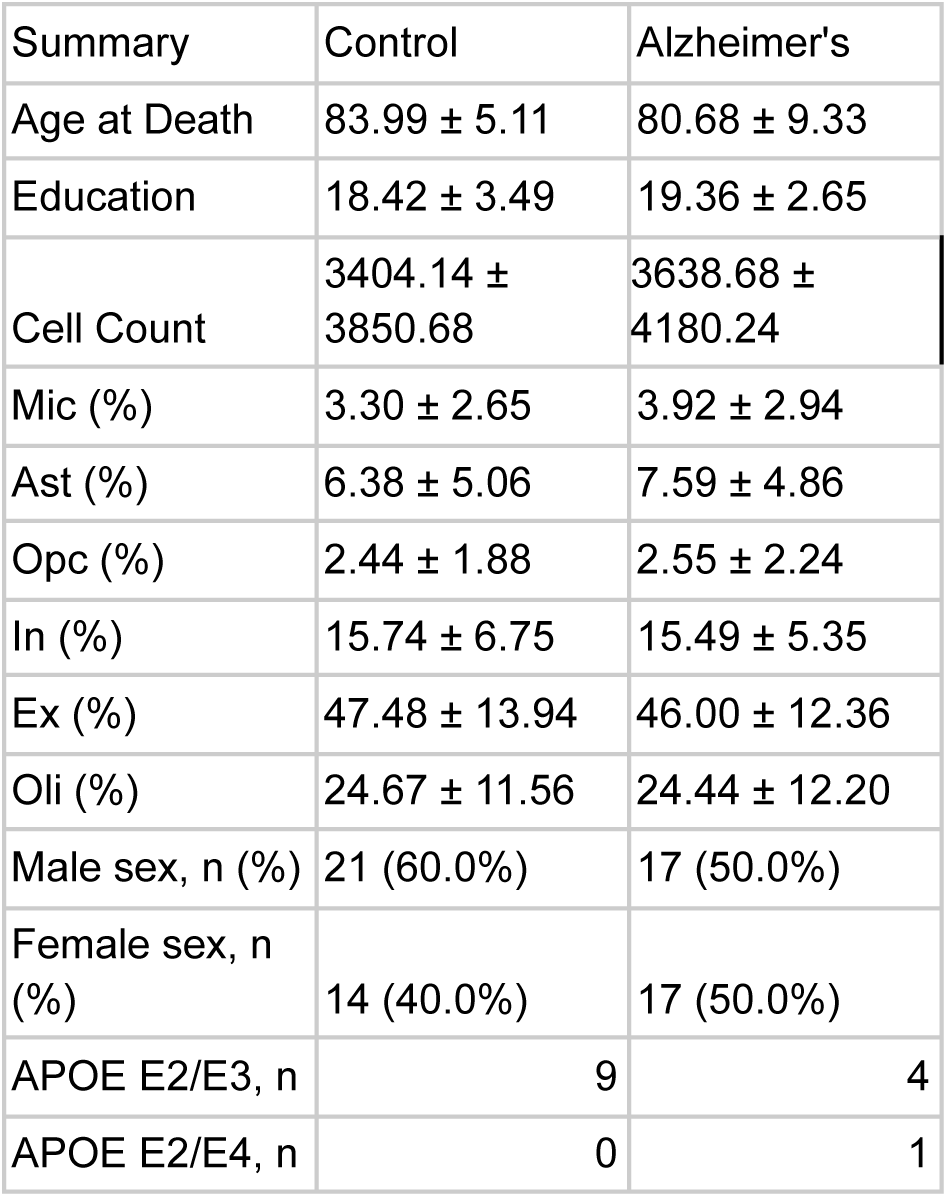
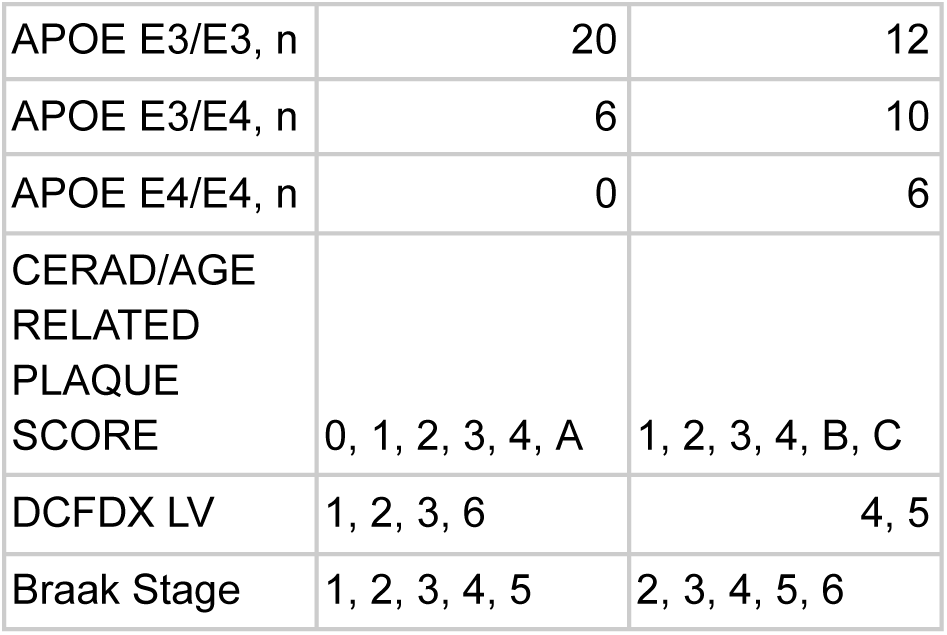
Summary statistics of all 69 donors used in our study.

To identify the sources of AD phenotypic heterogeneity across clinical and pathological definitions, approaches are required that jointly model gene expression with demographic and genetic variables to reveal multivariate signatures of AD phenotypes overlooked by standard DE analyses. To that end, TriSCOPE nominates disease-predictive genes overlooked by conventional analyses yet independently supported by genetic evidence, validated across cohorts, and anchored in brain regulatory architecture, and in doing so captures heterogeneity between phenotypic definitions. This integrative framework offers a new approach in the analytic toolbox for dissecting molecular risk across clinical symptoms, pathological burden, and disease progression in AD, establishing a framework for mechanistic dissection and therapeutic targeting.

## Results

### Predicting AD using Demographics, APOE genotype, and gene expression

We first deployed our pipeline, TriSCOPE, to identify molecular signatures of clinical Alzheimer’s disease (AD) that extend beyond known risk factors, developing an interpretable machine learning framework that prioritizes gene expression features predictive of clinical AD. Clinical AD was defined by the diagnosis rendered at the last visit while alive and was adjudicated by a panel of neurologists, neuropsychologists, and other clinicians. Our goal was to use machine learning to uncover combinations of demographic, genetic, and transcriptomic features that confer cellular vulnerability on a phenotype specific basis. We applied this approach to over 230,000 single-nucleus transcriptomes from two human postmortem PFC cohorts (70,634 cells in the discovery dataset; 172,514 cells in an independent validation set), integrating gene expression across 17,926 genes, APOE genotype, and demographic features including age at death, sex, and education (Fig. 1a).

**Figure 1:**
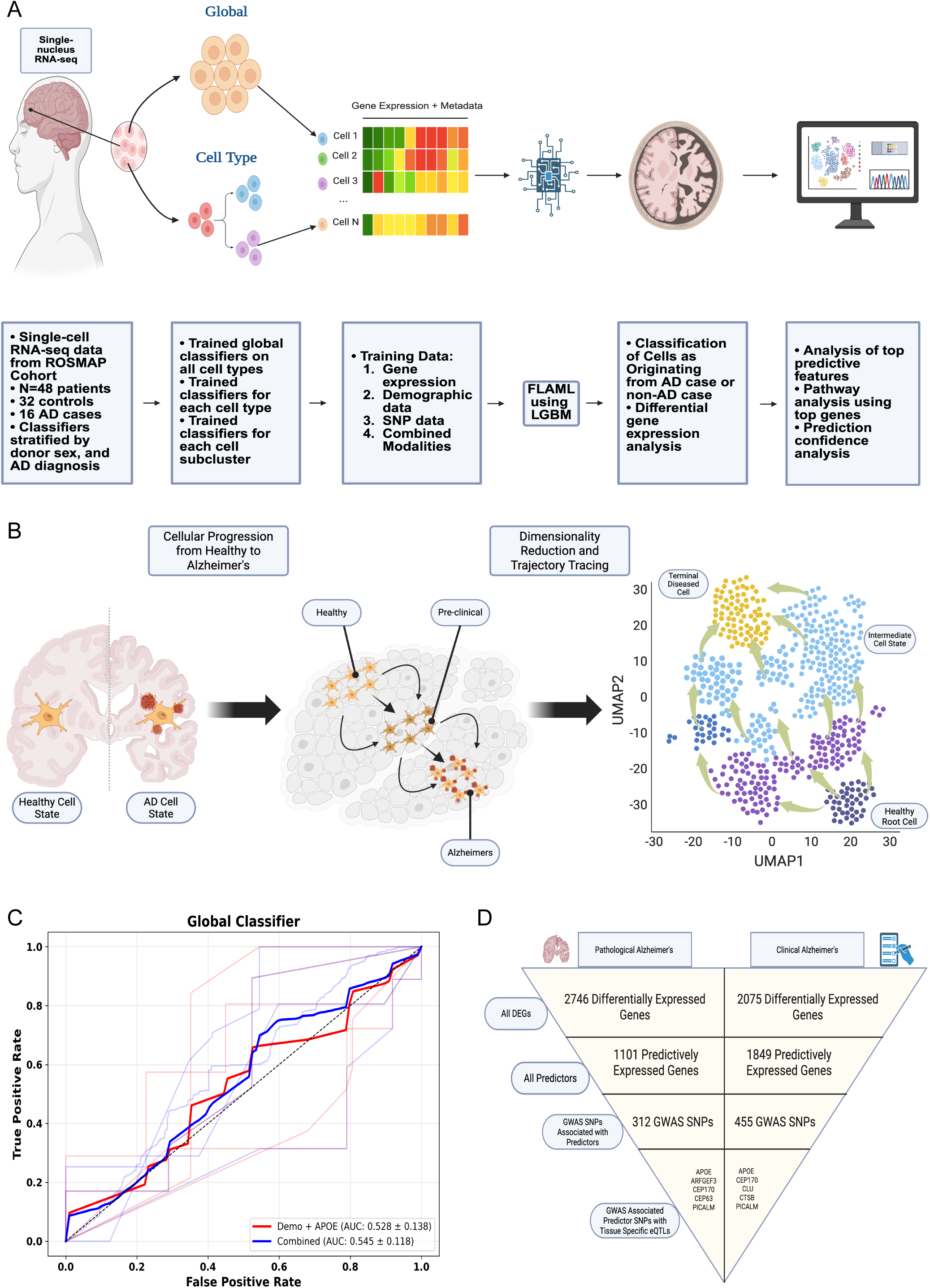
Overview of the TriSCOPE framework for integrative single-cell predictive and trajectory analysis in Alzheimer’s disease. a, Outline of the predictive modeling component of our TriSCOPE approach. Including cell-specific disease classification and downstream feature importance analysis. b, Outline of the pseudotime modeling component of TriScope. We choose cells based on healthy phenotype, no Amyloid pathology (CERAD), and high cognitive diagnosis scores (COGDX). Setting the transcriptional of this healthy cell as our root, we then trace the transcriptional progression away from this cell in order to map the progression of disease. We perform this at the cell subcluster level to avoid confounding with transcriptional differences between cell types. c, A predictive classifier trained across five splits on a global scale, no cell type differentiation, on Demographics and APOE data. The second curve shows a predictive classifier trained across five splits on a global scale, no cell type differentiation, on Demographics, APOE, and Gene Expression data. d, Pyramid shows how our predictive pipeline, both in isolation and when coupled with genetic anchoring can nominate a few high-confidence gene expression targets.

To this end, we first trained baseline models using only demographic features (age at death, sex, and years of education) and APOE genotype to predict clinical AD diagnosis. These models performed only slightly above chance (mean AUC = 0.528 ± 0.138), highlighting the limited explanatory power of demographics and APOE genotype at single-cell resolution (Fig. 1c). In contrast, incorporating gene expression features alongside demographics and APOE genotype modestly improved classification performance (mean AUC = 0.545 ± 0.118) (Table 1), suggesting that transcriptional profiles contain additional predictive information not captured by demographics and APOE genotype (Fig. 1d). This modest gain in predictive power motivated examining gene expression on a cell type specific basis as a lens into cellular states associated with disease, motivating deeper exploration of which genes contribute most to clinical AD classification across cell types.

### Cell Type-Specific Biomarker Discovery beyond Demographics and APOE genotype

To investigate how transcriptional vulnerability to Alzheimer’s disease varies across brain cell types, we trained separate models for each major population: Astrocytes, Excitatory Neurons, Inhibitory Neurons, Microglia, Oligodendrocytes, and Oligodendrocyte Progenitor Cells (OPCs). Specifically, within each cell type, we compared the performance of models with the following combinations of predictor variables 1) Demographics and APOE genotype, 2) Gene expression, and 3) the Combination of demographics, APOE genotype, and gene expression (Fig. 2a).

**Figure 2:**
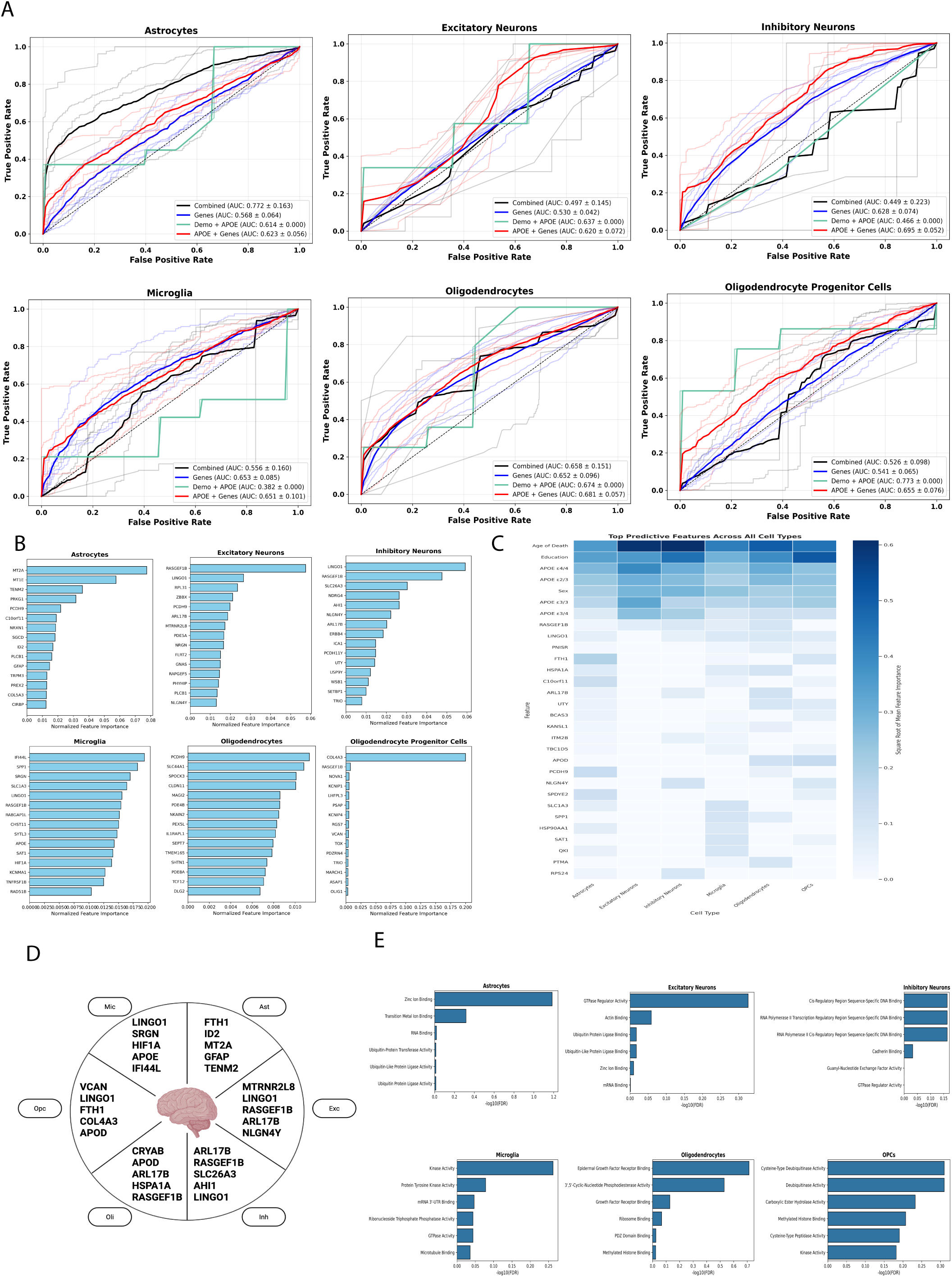
Cell-type-specific predictive modeling reveals diverse transcriptional determinants of clinical Alzheimer’s disease status. a, Receiver operating characteristic (ROC) curves summarizing the results from five independent runs of classification experiments performed on six major brain cell types: Astrocytes, Excitatory Neurons, Inhibitory Neurons, Microglia, Oligodendrocytes, and Oligodendrocyte Progenitor Cells. For each cell type, classifiers were trained using four feature sets: (1) a combined set including demographic variables (age at first diagnosis, sex), APOE genotype, and gene expression data (black curves); (2) demographic variables and APOE genotype only (grey curves); (3) gene expression plus APOE genotype (red curves); and (4) gene expression only (blue curves). Mean area under the curve (AUC) values ± standard deviation are reported in the figure legends for each curve. Lighter lines represent individual splits; bold lines indicate mean performance. Higher AUC values reflect better discrimination between Alzheimer’s disease and control cells. b, Top predictive genes identified from gene expression–based classifiers across major brain cell types. Normalized feature importance scores for the top 15 gene features used to classify Alzheimer’s disease versus control cells, shown separately for each major brain cell type: Astrocytes, Excitatory Neurons, Inhibitory Neurons, Microglia, Oligodendrocytes, and Oligodendrocyte Progenitor Cells. Feature importance was computed for each gene as the raw importance assigned by the classifier in each of five independent train-test splits, divided by the sum of all raw importances in that split to obtain a normalized score. Mean normalized importance across five splits was then calculated for each gene, and the 15 genes with the highest mean scores per cell type are displayed. Standard deviations are reported in Supplemental Table 1. Bars represent mean normalized importance, highlighting genes most strongly contributing to Alzheimer’s disease prediction in each cell type. c, Heatmap showing the top 30 most predictive features from the combined classifiers trained on demographic variables (age at death, sex, education), APOE genotype, and gene expression data, separately for each major cell type: Astrocytes, Excitatory Neurons, Inhibitory Neurons, Microglia, Oligodendrocytes, and Oligodendrocyte Progenitor Cells (OPCs). Feature importance for each feature was computed as the raw importance assigned by the classifier in each of five independent splits, normalized by the total importance within each split. The square root of the mean normalized importance across splits is shown for visualization. Rows represent features (genes and demographic variables), and columns represent cell types. Color intensity indicates relative feature importance, highlighting which features most strongly contributed to Alzheimer’s disease versus control classification in each cell type. d, Circle plot summarizing a selected set of key gene features identified from SHAP analysis of combined model classifiers (Supplemental Fig. 9a) and gene expression-based classifiers, chosen for their high normalized feature importance scores and biological relevance as potential Alzheimer’s disease predictors. Genes are shown for each major brain cell type: Microglia (Mic), Astrocytes (Ast), Excitatory Neurons (Exc), Inhibitory Neurons (Inh), Oligodendrocytes (Oli), and Oligodendrocyte Progenitor Cells (Opc). e, Bar plots showing top enriched biological pathways identified using pathway enrichment analysis of predictive genes (assigned predictive feature importance across two or more splits), performed separately for each major brain cell type: Astrocytes, Excitatory Neurons, Inhibitory Neurons, Microglia, Oligodendrocytes, and Oligodendrocyte Progenitor Cells. Enrichment was performed using g:Profiler, and odds ratios are shown on the x-axis, indicating the degree of overrepresentation of each pathway among nominated genes relative to the background gene set. Top terms highlight cell–type–specific processes potentially implicated in Alzheimer’s disease–associated transcriptional changes.

Our AD classifiers all demonstrated significant predictive performance across cell types (mean AUC: 0.620 ± 0.072 to 0.695 ± 0.052) using gene expression and APOE genotype alone relative to demographics and APOE genotype (Supplemental Table 4, Fig. 2a). To identify the molecular drivers underlying each model’s performance, we next ranked feature importance scores across all cell types, both in our combined data modality models and gene expression-only models. In our combined data modality model (3), age at death, years of education, APOE ε3/ε4 genotype, APOE ε4 homozygous genotype, sex, *ARL17B*, *HSPA1A*, *RASGEF1B*, and *LINGO1* were the most predictive features across cell types (Fig. 2c, d). These genes, which had high predictive importance across cell types after accounting for demographic and APOE genotype, may be reflective of global processes of Alzheimer’s disease. Among these globally predictive genes, *RASGEF1B* stood out as a member of the RAS signaling pathway, with an established role in synaptic plasticity and neuronal dysfunction ^19,20^. *LINGO1* is a known regulator of axonal regeneration and myelination, and has been implicated in Alzheimer’s disease ^21^. *ARL17B*, which encodes Rapidly Accelerated Fibrosarcoma (RAF) GTPases, likewise demonstrated pan-cell-type importance consistent with links between the GTP-binding protein complex and amyloid precursor protein (APP) to disease progression^22,23,24^. These findings reinforce that our model can uncover established drivers of Alzheimer’s disease. Still, we noted that pan-cell-type signals alone were insufficient to capture the full transcriptional heterogeneity of AD, motivating closer examination at the level of individual cell types.

While some features were shared across cell types, others were highly specific. Top predictors that were highlighted in only one cell type included (Supplemental Table 2) mitochondrial peptide genes in Excitatory Neurons^25^, chloride/bicarbonate exchanger, genes related to synaptic plasticity, and Y-chromosome linked genes in inhibitory neurons (Fig. 2b) (Supplemental Table 2)^26^. Unique predictors in microglia included immune activators (*IFI44L* ^27^, *SRGN, HIF1A, APOE*) ^28^ and *HSPA1A* (an inhibitor of amyloid-beta and tau neurofibrillary tangle formation) (Fig. 2b, Supplemental Table 2)^28,29^. In Oligodendrocytes, top predictive features included proteins linked to GFAP filament stabilization (*CRYAB* ^30,31^, *CLDN11*, and *APOD*) all of which were stronger predictors than APOE Genotype e3/e4 status (Fig. 2b; Supplemental Table 1). *COL4A3* emerged as a top predictor in OPCs (Fig. 2d; Supplemental Table 1; Supplemental Table 2). In Astrocytes, predictive features included *ID2*, involved in inflammatory signalling, and *FTH1* which is implicated in AD cognitive symptoms via Astrocyte Ferroptosis (*FTH1* ^32^), as well as the Astrocyte marker gene *GFAP* (Fig. 2b, Supplemental Table 2)^33^ ^34^.

The identification of well-characterized AD-related genes among top features reinforces the biological validity of our modeling approach, and suggests that those high-ranking predictors with limited prior characterization may also play meaningful roles in AD (Fig. 2e). Altogether, these findings nominate a spectrum of pan-cell-type and cell-type–specific biomarkers, providing a foundation for discovery of transcriptional drivers of AD.

### Differential expression identifies genes concordant and discordant with top cell-type specific predictors of AD

To contextualize and further interpret features identified by predictive modeling, we used the second branch of TriSCOPE to perform differential expression analysis between AD and control samples within each major cell type adjusting for APOE genotype and demographic covariates. In Excitatory Neurons, many of our top predictors such as *LINGOI, RASGEF1B*, and *MTRNR2L1* were significantly upregulated in AD while another predictor *MTRNR2L8* was downregulated (Fig. 3d) (Supplemental Table 8). While differential expression analysis revealed over 1,081 genes (FDR-corrected p-value < 0.05, abs(log2FC) > 0.25; Fig. 3b) altered in clinical AD within Excitatory Neurons, a much smaller subset of only 162 features (Fig. 4e) were predictive (24 overlapping, Fig. 3a) by our predictive modeling analyses, highlighting a much more focused core of disease-relevant changes when using a multivariate machine learning approach as opposed to a univariate differential expression approach. Similarly in Microglia, several top predictive genes were also significantly differentially expressed including *IFI44L*, the highest-ranking predictor, was significantly downregulated (log₂FC = –1.21, FDR = 0.0006), consistent with prior evidence implicating impaired type I interferon signaling, and reduced expression of interferon-stimulated genes like *IFI44L*, in individuals at elevated risk of AD progression ^35^ (Fig. 3c). *SRGN* and *HSPA1A*, associated with immune response and protein quality control were also downregulated while *APOE* (log₂FC = 0.36, FDR = 0.009) was upregulated (Fig. 3c, Supplemental Table 8). Oligodendrocytes had the largest predictor-DEG overlap with 102 genes.

**Figure 3:**
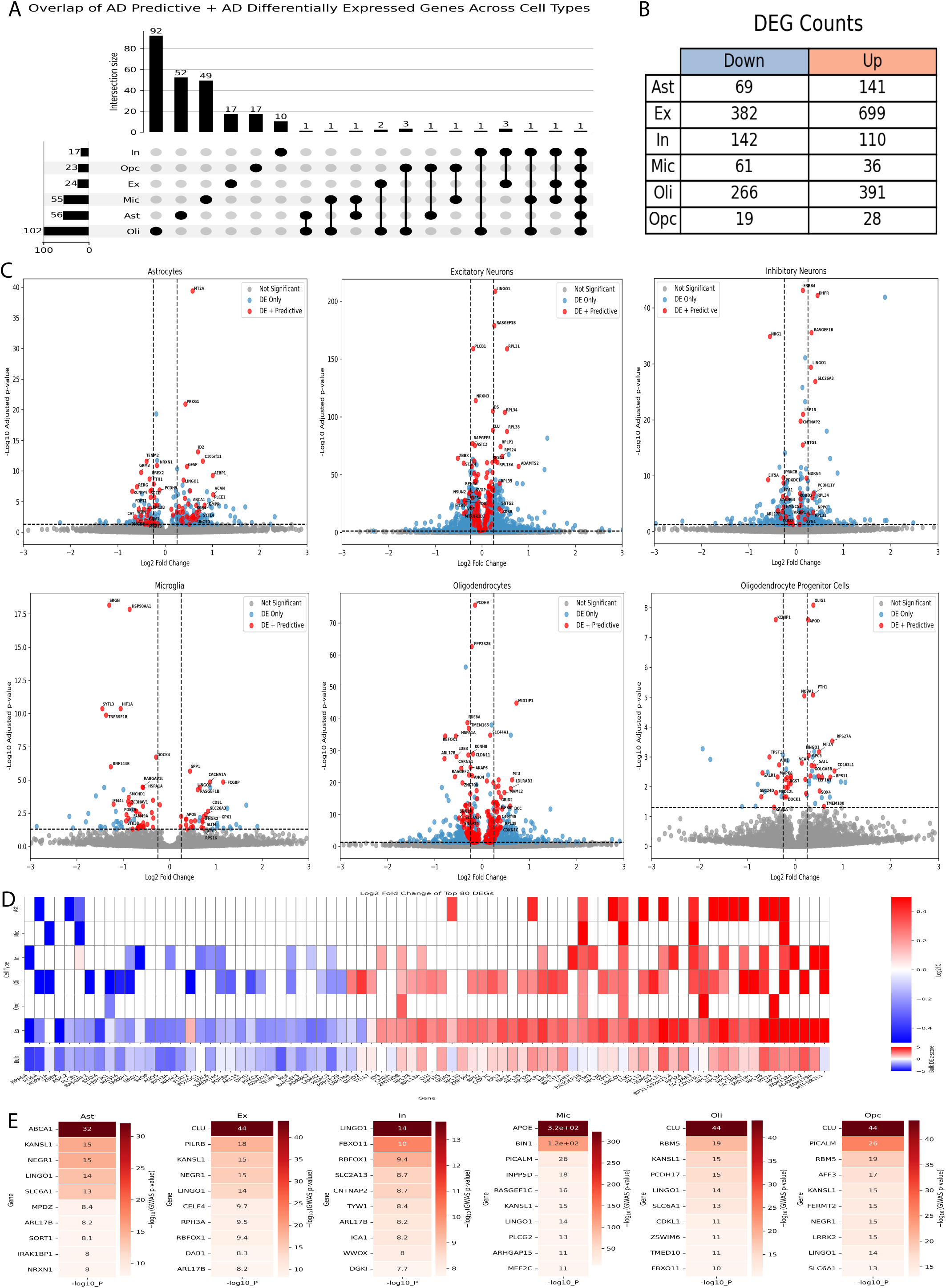
Integration of predictive and differential expression signatures identifies convergent disease markers across cell types. a, Upset plot illustrating the overlap between genes identified as predictive (assigned nonzero feature importance in at least two out of five independent classifier splits) and genes identified as differentially expressed (FDR-adjusted *P* < 0.05) in Alzheimer’s disease versus control cells, analyzed separately for each major brain cell type: Astrocytes (Ast), Excitatory Neurons (Ex), Inhibitory Neurons (In), Microglia (Mic), Oligodendrocytes (Oli), and Oligodendrocyte Progenitor Cells (Opc). Bar heights represent intersection sizes, and connected dots indicate specific combinations of cell types sharing overlapping genes. Notably, the strongest overlap was observed in astrocytes and excitatory neurons, suggesting a substantial convergence of predictive and transcriptionally dysregulated signals in these cell types. b, Table summarizing the number of differentially expressed genes (DEGs) identified as significantly downregulated (blue column) or upregulated (orange column) in Alzheimer’s disease versus control cells across each major brain cell type. Differential expression was performed using a Poisson generalized linear mixed model, including an offset for library size and controlling for post-mortem interval, age at death, and sex (FDR-adjusted *P* < 0.05). c, Volcano plots showing differentially expressed genes (DEGs) in Alzheimer’s disease versus control cells for each major brain cell type: Astrocytes, Excitatory Neurons, Inhibitory Neurons, Microglia, Oligodendrocytes, and Oligodendrocyte Progenitor Cells. Genes are plotted by log2 fold change (x-axis) and –log10 FDR-adjusted *P*-value (y-axis). Genes significant for differential expression (FDR-adjusted *P* < 0.05) and also identified as predictive (nonzero feature importance in at least two splits) are highlighted in red. Genes significant for differential expression only are shown in blue, and non-significant genes are shown in grey. Selected predictive DEGs are labeled, illustrating key markers that contribute to both differential transcriptional changes and disease-state classification within each cell type. d, Heatmap displaying log2 fold changes of the top 80 differentially expressed genes (DEGs) in Alzheimer’s disease versus control cells across six major brain cell types. Genes were selected based on having an absolute log2 fold change > 0.2 in at least one cell type and were ranked by minimal FDR-adjusted *P*-value across all cell types. Color intensity reflects log2 fold change, with red indicating upregulation and blue indicating downregulation in Alzheimer’s disease. Only genes with FDR-adjusted *P* < 0.05 are shown. e, Heatmaps showing –log₁₀transformed GWAS *P*-values for top genes overlapping between predictive genes identified in each cell type and Alzheimer’s disease genome-wide association study (GWAS) loci (P < 5 × 10⁻⁸). Up to 10 most significant genes are shown per cell type. Color intensity reflects GWAS significance, with darker red indicating stronger genetic association. Notably, APOE exhibited an extremely strong GWAS association in microglia.

**Figure 4:**
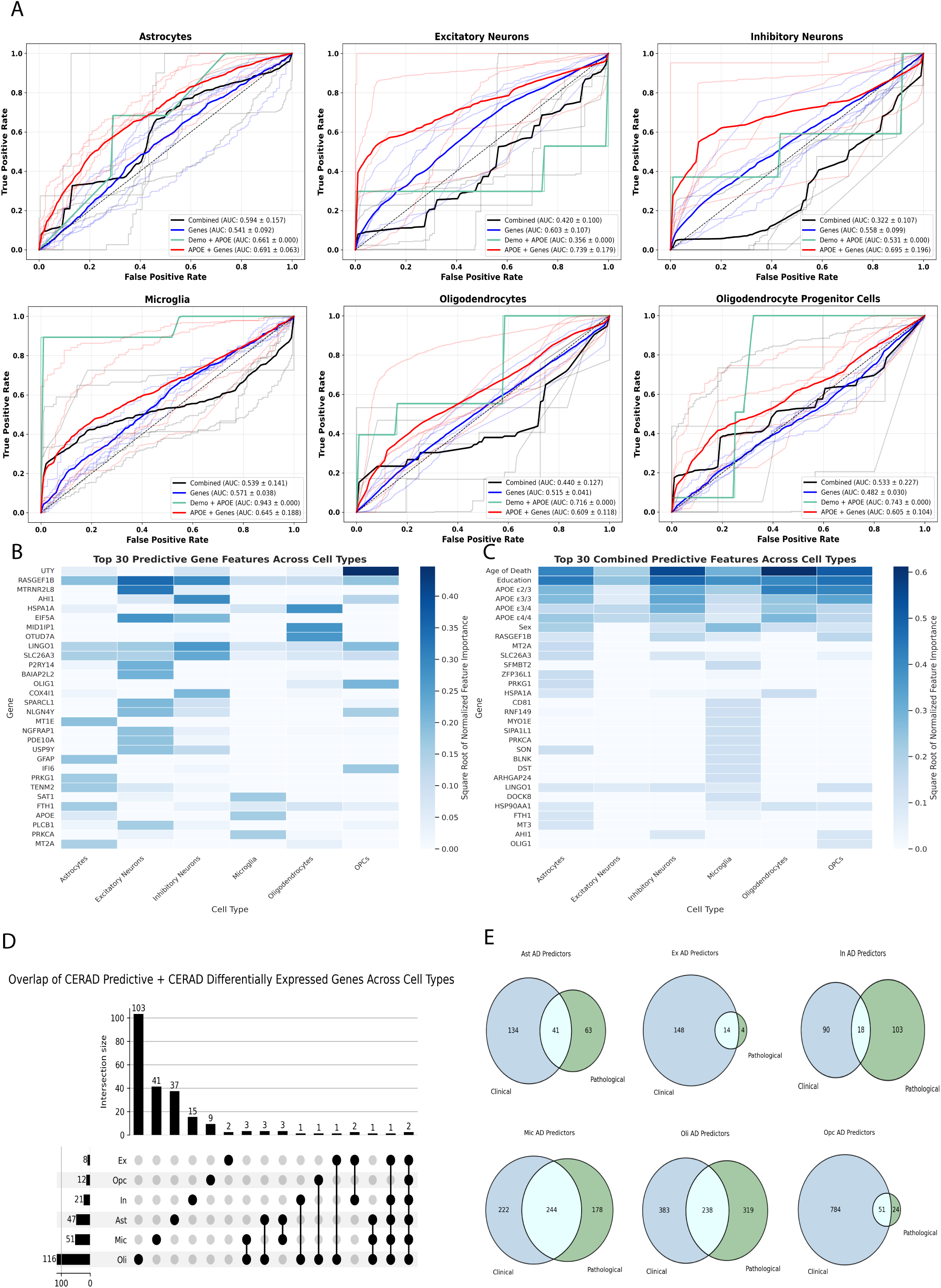
Predictive modeling of neuropathologic amyloid burden uncovers distinct and overlapping molecular signatures across cell types. a, Receiver operating characteristic (ROC) curves summarizing the results from five independent runs of classification experiments performed on six major brain cell types: Astrocytes, Excitatory Neurons, Inhibitory Neurons, Microglia, Oligodendrocytes, and Oligodendrocyte Progenitor Cells. In this analysis, the classification task was to distinguish cells from individuals with high Alzheimer’s pathology (CERAD score 1 or 2) versus controls. For each cell type, classifiers were trained using four feature sets: (1) a combined set including demographic variables (age at first diagnosis, sex), APOE genotype, and gene expression data (black curves); (2) demographic variables and APOE genotype only (red curves); (3) gene expression plus APOE genotype (green curves); and (4) gene expression only (blue curves). Mean area under the curve (AUC) values ± standard deviation are reported in the figure legends for each curve. Lighter lines represent individual splits; bold lines indicate mean performance. Higher AUC values reflect better discrimination of cells from high- versus low-pathology individuals. b, Heatmap showing the relative importance (squared root-normalized feature importance) of the top 30 most predictive gene expression features identified for CERAD score classification, across each cell type. Darker blue indicates higher importance. Gene features were ranked by their mean importance across cross-validation splits and then displayed in order of aggregate contribution. Feature importance was square-root-normalized for display. c, Heatmap showing the relative importance of the top 30 combined features (genes, APOE genotype, and demographic features) for CERAD score classification, across each cell type. This includes non-gene features (e.g., APOE ε2/ε3/ε4 status, age at death), highlighting their potential contributions relative to gene expression markers. Feature importance was squared root-normalized for display. d, UpSet plot illustrating the overlap between the top predictive genes for CERAD score classification and differentially expressed genes (DEGs) identified between CERAD high and low pathology groups within each cell type. Vertical bars indicate intersection sizes (overlap), and connected dots indicate cell types contributing to each intersection set. This analysis highlights cell-type-specific overlaps and shared predictive signatures. e, Venn diagrams summarizing the overlap between gene features predictive of CERAD score pathology status (blue) and gene features predictive of Alzheimer’s disease diagnosis (green), for each major cell type. Gene predictors were defined as features that appeared in at least two out of five cross-validation splits. The overlap highlights shared versus distinct molecular predictors used to discriminate pathology severity (CERAD) and clinical AD status across cell types.

Overall, we found varying overlap between our gene-expression-based predictors and DEGs (2.75-32% of predictive genes were DEGs across cell types), highlighting the capacity of multivariate modeling to uncover disease-relevant features that may not exhibit strong univariate shifts but could participate in predictive expression patterns in AD.

### Clinical and Neuropathological Predictors Diverge Across Cell Types

Given AD’s two hallmarks, cognitive decline and ATN (amyloid, tau, and neurodegenerative) pathology, we suspected that gene expression and other biological features may explain AD neuropathology (i.e., Consortium to Establish a Registry for Alzheimer’s Disease (CERAD) score) differentially from clinical AD diagnosis adjudicated by a panel of neurologists, neuropsychologists, and other clinicians. 20/48 donors had neither CERAD pathology or Clinical AD diagnosis, 4 were diagnosed with AD but did not meet the CERAD pathology criterion, 6 met the CERAD pathology criterion but were not clinically diagnosed, and finally 18 met both criteria (Supplemental Fig. 3a). Thus, our second goal was to determine whether gene expression could reveal cell-type specific signatures of Alzheimer’s disease phenotypes beyond what is captured by known demographic and genetic risk factors.

To test this, we reran our analysis of the Mathys et al. dataset^7^, redefining outcomes by neuropathology rather than clinical diagnosis. Specifically, we classified moderate or frequent neuritic plaques (CERAD score 1-2) as AD, using this semiquantitative measure of plaque pathology as the outcome. Our classifiers showed that gene expression together with APOE genotype strongly predicted neuritic plaque pathology across cell types, similar to the clinical diagnosis of AD (Fig. 4a).

Across all cell types, APOE genotype and gene expression predicted neuritic plaque pathology (mean AUC range: 0.551 (Oligodendrocytes)-0.739 (Excitatory neurons) Supplemental Table 16). Across cell types, we found strong concordance between genes predictive of pathology AD and clinical AD (Fisher’s q < 1.92e-20) (Supplemental Table 7), but especially in Microglia with 244 overlapping predictors (OR = 106.7, 95% CI: 84.4-134.9) and in Oligodendrocytes 238 overlapping predictors (OR = 33.09, 95% CI: 27.2-40.27) (Fig. 4e, Supplemental Table 7). These results suggest in Microglia and Oligodendrocytes, molecular signals of amyloid pathology may be tightly coupled to clinical manifestation of disease ^36,37^.

Although many predictors overlapped with clinical AD, their relative importance shifted in the pathology-based models. *RASGEF1B* once again emerged as a pan-cell type predictor (Fig. 4b, Supplemental Table 3). In Microglia, top predictors included *SAT1*, *PRKCA*, and *APOE*. Both *SAT1* and *PRKCA* were downregulated in AD, complementing our prior observation of reduced interferon signaling, while *APOE* was upregulated, consistent with advanced amyloid pathology (Fig. 4b–c, Supplemental Table 3, 7) ^38^. Interestingly, the pathology-based models also highlighted genes that did not predict clinical AD, suggesting distinct molecular drivers of plaque pathology. For example, in Astrocytes we again observed *RASGEF1B* and *GFAP*, but APOE expression emerged in addition as a strong predictor. In contrast, *CLU,* while predictive of clinical AD, was not predictive of neuropathology, suggesting a potential bias toward cognitive AD phenotypes. Together, these findings reveal that while clinical AD and AD neuropathology share many overlapping molecular programs, their divergence across specific genes highlights distinct biological processes underlying cognitive decline versus amyloid pathology in a cell type-specific manner.

### Genetic Risk Loci Map to Cell-Type–Specific Predictors of clinical AD and AD neuropathology

We next asked whether our expression-based predictors, derived separately for clinical AD and AD neuropathology, were supported by genome-wide significant AD loci. Using the NHGRI-EBI GWAS Catalog ^39^, we tested whether the union of gene expression predictors for clinical AD and AD neuropathology were proximal to previously reported AD-associated SNPs, and further assessed whether these SNPs act as cis-eQTLs in the brain.

Clinical AD predictors in Inhibitory Neurons, Microglia, Oligodendrocyte Progenitor Cells, and Oligodendrocytes were significantly enriched for genes containing genome-wide significant SNPs (Fisher’s p range=1.78e-5 - 0.047). In Oligodendrocytes, OPCs, and Excitatory Neurons, the predictive gene with the strongest genetic association with AD was *CLU* (beta=78.65, -log10p=32), and *CLU* gene expression levels have been implicated in cognitive decline in Alzheimer’s ^40^ (Fig. 3e, Supplemental Table 5). Microglia showed 68 predictive overlapping GWAS genes (OR = 1.48, Fisher’s p = 0.047), including *APOE*, *BIN1*, *PICALM*, and *RASGEF1C* (Fig.3e, Supplemental Table 5). ABCA1 was the top GWAS SNP in Astrocytes (-log10p = 32) (Fig. 3e).

To further examine functional relevance, we assessed whether these AD-associated SNPs residing in cell-type-specific predictive genes were also cis-expression quantitative trait loci (eQTLs) using the GTEx brain tissue eQTL databases ^41^. We found that across all cell types, 5 SNPs were both eQTLs in the frontal cortex and associated with AD in GWAS (Supplemental Fig. 5d). Four of these resided in *APOE*, *CEP170*, *CTSB*, and *PICALM*, all predictors unique to Microglia, while *CLU* was predictive in Oligodendrocytes and Excitatory Neurons. *CLU* and *PICALM* were predictive in Oligodendrocyte Progenitor Cells, hinting at a role for OPCs clearance of β-amyloid ^42^.

We repeated this overlap analysis using predictors of AD neuropathology rather than clinical diagnosis, which revealed *APOE* as containg the top SNP in both Astrocytes and Microglia (Supplemental Fig. 6c, Supplemental Table 6). We once again found the strongest overlap with GWAS hits of 107 predictive genes in Oligodendrocytes (OR = 1.85, Fisher’s p = 1.27E-07) (Supplemental Table 6). We then took these GWAS associated neuropathology predictors and checked for brain tissue cis-eQTLs, revealing 5 SNPs. These included *APOE* in Astrocytes, *APOE* and *PICALM* in Microglia, *CEP170* and *CEP63* in Inhibitory Neurons, and *ARFGEF3* in OPCs. By linking these SNPs to gene expression, and gene expression to AD, we establish a functionally interpretable link between these previously GWAS associated and gene expression that is predictive of Alzheimer’s disease (Fig. 1d). This genetic anchoring not only validates the biological relevance of our predictors but also reveals phenotype-specific differences, suggesting that distinct regulatory programs may operate at different stages or manifestations of AD.

### Cell-type-specific expression differences among AD neuropathology-positive patients with and without clinical AD

To better understand the gene expression changes associated with clinical AD when AD neuropathology is present, we performed differential expression analysis between the 6 individuals who exhibited Alzheimer’s pathology but without clinical diagnosis, and the 18 with Alzheimer’s pathology *and* clinical diagnosis. Our analysis revealed an upregulation of *SRGN*, *HIF1A*, and *RASGEF1C* in Microglia, and subsequent pathway analysis revealed enrichment of pathways related to neuroinflammatory response, cytokine production and interleukin-6 regulation (Supplementary Fig. 2a, b). Similar analysis in Oligodendrocytes revealed upregulation of pathways related to cellular response to unfolded and topologically incorrect protein and pathways related to long-term memory (Supplemental Fig. 2c). These results suggest that within individuals with equivalent neuropathological burden, additional microglial and oligodendrocytic transcriptional programs may differentiate who progresses to clinically symptomatic AD.

### Predictive and differentially expressed genes replicate across cohorts and cell types

To confirm the robustness of these findings, we validated our cell-type-specific classifiers on an independent dataset comprising 12 clinical AD and 9 clinical control individuals, covering 169,496 nuclei across Microglia, Oligodendrocytes, Excitatory Neurons, Inhibitory Neurons, and Astrocytes (Supplementary Table 9). Using one-shot classifiers from our original dataset, we showed consistent predictive performance across cell types suggesting the generalizability of our models (Supplementary Fig. 1a). Gene-only one shot classifiers showed mean AUCs ranging from 0.56 (Microglia) - 0.67 (Astrocytes) (Supplementary Fig. 1b).

To assess the concordance of feature importance in both datasets, we fit autoML models from scratch on our validation dataset. We observed strong concordance in top predictive genes, once again with cell type specificity (Supplementary Fig. 1a; Supplemental Table 10). These included *NLGN4Y* in Inhibitory Neurons and *MTRNR2L8* in Excitatory Neurons (Supplementary Fig. 1c, Supplemental Table 10). Microglia showed *PLXDC2* and *APOE* as top predictors and DEGs (Supplemental Fig. 1d), while Astrocytes showed *COL21A1* and *GFAP*. In Oligodendrocytes we observed *CRYAB*, *FTH1*, and *APOD* to all be top predictors. We once again saw *RASGEF1B*, *ARL17B*, *LINGO1*, and *SLC26A3* as predictive across multiple cell types, reinforcing their robustness as pan-cell-type AD predictors (Supplemental Table 11, Supplemental Fig. 1a). We found strong overlap in predictive genes (FDR adjusted Fisher’s p < 0.05 in all cell types) (Supplemental Fig. 1a, Supplemental Table 11), including 40 overlapping predictors in Astrocytes (OR = 30.28) and 106 overlapping in Oligodendrocytes (OR = 21.78) (Supplemental Fig. 1a, Supplemental Table 11). We then measured differential expression concordance betweewn cell types and found highly significant DEG overlap across all 5 major cell types (OR: 2.88 to 7.50; FDR adjusted Fisher’s p < 3.02E-19 in all cell types; Supplemental Table 11). Overall, these replication analyses underscore the generalizability and robustness of our predictors across independent cohorts and reinforce their potential as reliable cell-type specific biomarkers of AD.

### Subcluster Level Expression Improves AD Prediction

After discovering cell-type specific genes both predictive and differentially expressed in clinically defined AD, we examined whether gene expression could also predict clinical AD on a cell subcluster level. To discover transcriptional subclusters that may provide accurate snapshots of disease, we performed gene expression-based prediction at the cell-type subcluster level. This analysis revealed Inhibitory 5 (In5) (top 3 marker genes: *CPLX3*, *TRPC3*, *KIT*) (mean AUC = 0.77 ± 0.079), Excitatory Neurons 6 (top 3 marker genes: *CA4*, *TIMP3*, *P2RY14*) (mean AUC = 0.704 ± 0.143), and Microglia 1 (top 3 marker genes: *RPL19*, *RPS11*, *RPL35*) (mean AUC = 0.669 ± 0.076) (Fig. 5a) as the most predictive cell type subclusters. At the subcluster resolution we saw that gene expression could predict clinical disease status in 31 out of 41 subclusters (mean AUC > 0.5), highlighting that localized gene expression patterns may reflect AD vulnerability (Fig. 5a, Supplemental Table 14). In Inhibitory Neurons 5, the top features were *RASGEF1B* and *PNISR* (Supplemental Table 10). We noted *LRRTM4*, which encodes leucine-rich repeat transmembrane protein 4, and *ATPB1* as top predictors in Ex6 (Supplemental Table 13). Taken together, these findings show that subcluster-level analyses can uncover transcriptional gradients overlooked at the broad cell-type resolution, and that focusing on these finer-grained states yields improved predictive performance, suggesting that disease vulnerability is better captured at the resolution of discrete transcriptional subclusters rather than whole cell types.

**Figure 5:**
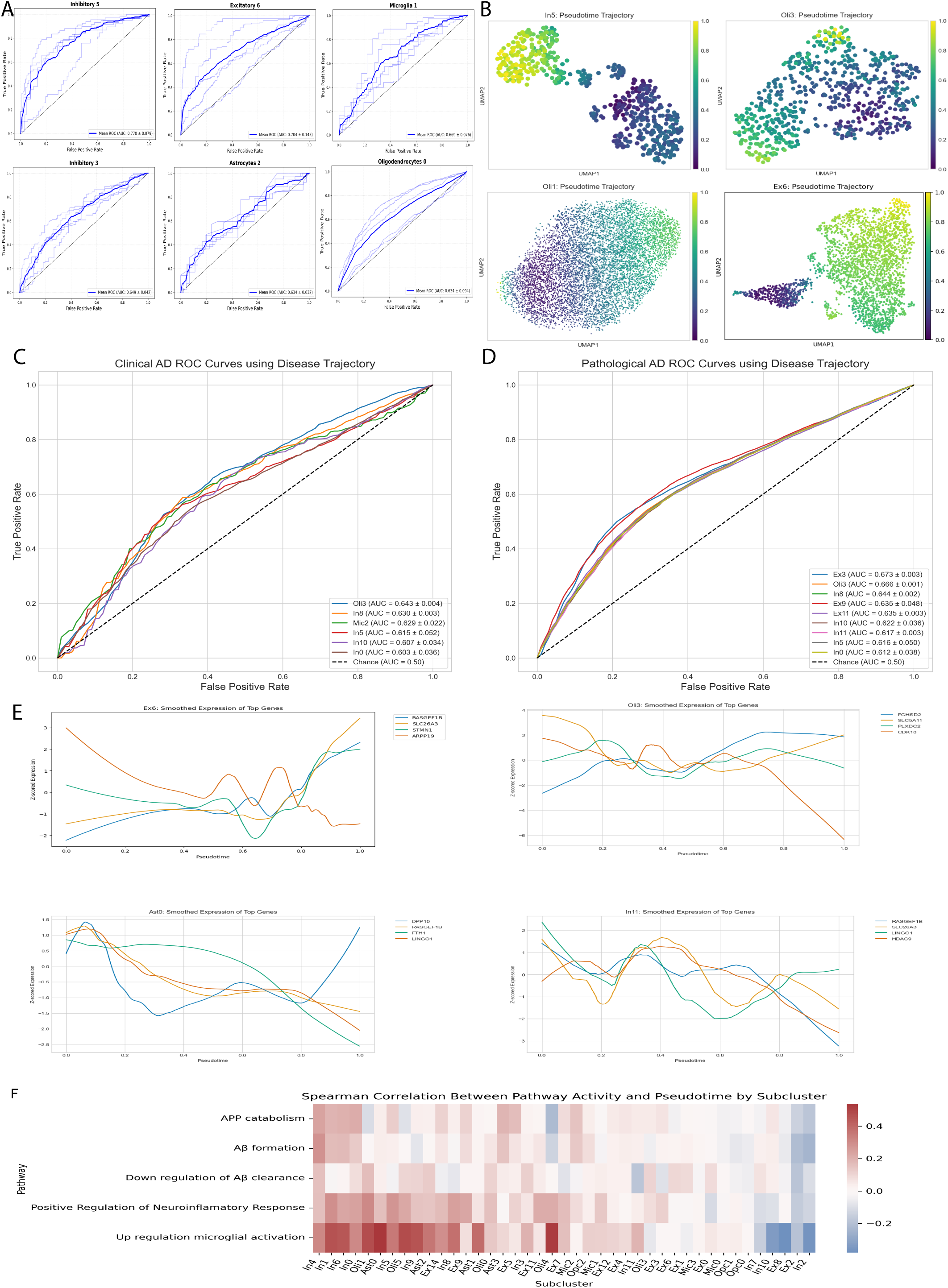
Subcluster-level predictive, trajectory, and trait enrichment analyses uncover fine-grained transcriptional programs in Alzheimer’s disease. a, Receiver operating characteristic (ROC) curves summarizing subcluster-level classification of Alzheimer’s disease status using gene expression data alone. The six subclusters that exhibited the highest mean ROC performance among all subclusters analyzed. Mean area under the curve (AUC) values ± standard deviation across five cross-validation splits are indicated in the figure legends. Light blue lines represent ROC curves from individual splits, and bold lines indicate the averaged ROC curve per subcluster. Higher AUC values reflect better discrimination of Alzheimer’s disease cells from control cells within each subcluster. b, Pseudotime trajectory visualizations for eight representative subclusters with high predictive performance or biologically relevant transcriptional signatures. For each subcluster, pseudotime was inferred using diffusion pseudotime (DPT), starting from root cells selected based on advanced amyloid pathology but minimal cognitive decline. Cells are plotted in UMAP space and colored by pseudotime values (color scale: dark purple to bright yellow), representing transcriptional progression from early to advanced disease states. c, ROC curves summarizing the predictive capacity of pseudotime trajectories for distinguishing clinical AD versus control cells across significant subclusters. Pseudotime was inferred using diffusion pseudotime (DPT) initialized from root cells with a low amyloid pathology score and low cognitive impairment. Shown curves represent subclusters with robust mean AUC values exceeding 0.60. d, ROC curves summarizing the predictive capacity of pseudotime trajectories for distinguishing pathological AD versus control cells across significant subclusters. d, Smoothed expression trajectories of top trajectory-influencing genes across pseudotime in subclusters Excitatory 6, Oligodendrocytes 3, Astrocytes 0, and Inhibitory Neurons 11. e, Heatmap showing Spearman correlation coefficients between pathway activity scores for five key Alzheimer’s linked biological processes (APP catabolism, amyloid-β formation, negative regulation of amyloid-β clearance, positive regulation of neuroinflammatory response, and positive regulation of microglial activation) and diffusion pseudotime values across all 41 subclusters.

### Subcluster Profiles Reveal Transcriptional Gradients of AD Progression

To further map the transcriptional axis of both clinical and pathological manifestations of AD within in subclusters, we performed diffusion pseudotime analysis^43,44^ by selecting a “healthy” root cell, based on no presence of AD neuropathology and no clinical AD diagnosis, and mapping the transcriptional progression away from this root cell based only on DEGs (Fig. 1b). In the 41 subclusters identified by Mathys et al ^7^, our pseudotime trajectory inference created a continuity between cells based on disease state. Cell pseudotime was predictive of AD neuropathology in 18 subclusters (mean AUC from 0.55-0.67), suggesting that gene expression within subclusters of these cells provides a predictive snapshot of Alzheimer’s neuropathology (Fig. 5b, Supplemental Table 14).

In Oligodendrocyte subcluster 3 (top marker genes: *HSPA1B*, *HSPA1A*, *HSP90AA1*), and Inhibitory Neurons 5 and 8 (*CHST9*, *PDGFD*, *CA1*), pseudotime values predicted clinical AD in a logistic regression model (mean AUC = 0.615 (± 0.052) to 0.643 (± 0.004); OR of 4.02 to 9.19; FDR-adjusted q = 1.8E-05 to 1.24E-72) (Fig. 5b-e; Supplemental Table 14). Across 41 studied subclusters, we found *RASGEF1B* to be a top trajectory influencing gene in 19 subclusters, with *LINGO1* and *SLC26A3* being top trajectory influencers in 16 subclusters (Fig. 5d). Additionally, there were distinct cell-type-specific genes influencing trajectories, such as *GFAP* in Astrocytes and *PLXDC2* in Oligodendrocytes (Supplementary Table 8), concordant with our predictive modeling analysis.

We then confirmed later pseudotime values mapped to transcriptional programs related to Alzheimer’s disease progression ^45^. For five amyloid-related Gene Ontology terms, we found 27 out of 41 subclusters showed a positive correlation between pseudotime values and APP catabolism and Amyloid beta formation, while 31 subclusters showed a positive correlation between pseudotime values and positive regulation of Microglia cell activation (Fig. 5e). Our trajectory analyses reveal that AD progression is not uniform but follows subcluster-specific transcriptional gradients, nominating key genes as potential markers of disease state and vulnerability.

## Discussion

Using single-cell gene expression, AD-associated variants, and machine learning, differential expression, and pseudotime analysis, we uncovered convergent gene expression programs that predict clinical AD diagnosis and AD neuropathology in a cell type-subcluster-specific manner. Across six major brain cell types, we systematically mapped Alzheimer’s disease (AD) vulnerability at both cell-type and subcluster resolutions independent of demographics and APOE. A central strength of our TriSCOPE framework emerged as its ability to identify a more concise, high-confidence set of disease-relevant features than traditional differential expression analysis, including genes that would not pass conventional univariate thresholds. This approach recovered well-established AD genes alongside underappreciated candidates such as *ARL17B* across cell types and *IFI44L* in Microglia—highlighting novel molecular features of disease vulnerability.

We identify *ARL17B* as a consistent pan-cell predictor of AD, aligning with recent evidence that the 17q21.31 MAPT inversion region exerts broad regulatory effects on brain phenotypes ^46^. Although *ARL17B* itself is poorly characterized, it tracks with the inversion haplotype that also modulates KANSL1 and LRRC37A, genes repeatedly implicated in neurodegeneration and AD GWASs ^47^. Zhukovsky et al. similarly linked *ARL17B* expression in frontal cortex to genetic risk and cortical thickness, underscoring its role as a haplotype-anchored marker of disease vulnerability ^48^. Notably, *ARL17B* emerged in our models after adjusting for APOE genotype, sex, and age, suggesting it represents a robust, covariate-independent marker of disease risk (and why it may have been overlooked in previous studies) and reproduced in an independent cohort. As such, *ARL17B* highlights the 17q21.31 locus as a genetically anchored, and part of a cell-wide regulatory hub, making it a compelling point for mechanistic studies. On the other hand, we also highlight IFI44L as a microglia-specific predictor of AD, reflecting its role as an interferon-stimulated gene and marker of innate immune activation. Unlike pan-cell predictors, *IFI44L* points to a cell-type–restricted program of interferon-driven microglial dysfunction.

Our findings build on, but are distinct from, important contributions of data and approaches from others. For example, whereas Mathys and colleagues provided essential foundational maps of cell-type and subcluster transcriptional profiles in Alzheimer’s disease ^7^, our work leverages an integrative and multifeature framework (TriSCOPE) to (a) uncover not only cell-specific but also convergent pan-cell-type predictors of disease that persists after rigorous adjustment for demographic and genetic covariates, (b) link molecular programs to both clinical and pathological definitions of AD, and (c) anchor transcriptomic changes in genetic risk loci—representing an advance in analytic precision, biological insight, and potential translational relevance over prior approaches.

Our analysis also uncovered heterogeneity between predictors of AD neuropathology and clinical AD, which is especially significant given that many current studies choose phenotypes that are a mix of clinical and pathological definitions ^49^, missing the opportunity to identify sources of clinical versus pathological variation. Across cell types, there were overlapping and distinct sets of predictors. Microglia and Oligodendrocytes showed the largest overlap between clinical and pathology-based predictors, suggesting close integration of neuroinflammatory and axonal myelination maintenance pathways with both cognitive decline and neuropathological burden.

Integration with GWAS and brain eQTL datasets provided support for many predictors, strengthening their biological evidence. One-to-one SNP-to-gene mapping allowed direct alignment of predictive genes with genome-wide significant AD loci. In microglia, *APOE* and *PICALM* overlapped with pathology-associated predictors, pointing to amyloid clearance and plaque-associated immune pathways. Clinical predictors in Oligodendrocyte Progenitor Cells included CLU, implicated in amyloid processing and cognitive decline, while astrocyte predictors included ABCA1, a lipid transporter linked to cognitive deficits in mice models ^50^. These cell-type-specific overlaps link genetic susceptibility loci to transcriptional states in a biologically interpretable way, nominating high-confidence targets for mechanistic follow-up. Finally, pseudotime analysis at the subcluster level revealed progressive transcriptional trajectories predictive of AD. Key genes such as *RASGEF1B*, *SLC26A3*, and *LINGO1* defined the disease trajectory across multiple subclusters validating their significance, and pseudotime states were positively associated with amyloid formation, impaired clearance, and neuroinflammatory pathways, highlighting transcriptional progression within discrete neuronal and glial subpopulations and suggesting potential key markers of vulnerability ^51^.

Our findings also have translational implications for therapeutic development. Several of the top predictors we identified—such as *LINGO1*, *RASGEF1B*, and *CRYAB*—have either been directly investigated or are biologically tractable drug targets. For example, *LINGO1* inhibitors were previously tested in phase II multiple sclerosis trials as potential remyelination-promoting agents ^52,53^. Preclinical work has also shown that hippocampal overexpression of *Lingo1* improves cognitive function in a mouse model of AD, with knockdown showing the opposite effect ^52,53^. Similarly, *RASGEF1B* has been implicated in inflammatory signaling, and its regulation of Intercellular Adhesion Molecule 1 suggests that modulating this axis may influence neuroinflammatory cascades in AD ^54^. Additionally, *CRYAB* protein has previously been identified as a myelin binding partner and linked to increased phagocytosis of the myelin sheath ^55^. Together, these results highlight genetically anchored, cell-type–specific molecular programs that may guide targeted therapeutic development in Alzheimer’s disease, and demonstrate how our pipeline can systematically nominate such high-confidence targets.

Our framework is predictive in design, but its primary aim is feature discovery rather than clinical prognostication. The AUCs reflect the use of post-mortem, cross-sectional data, where prediction cannot be the endpoint; instead, the value lies in identification of key genes and cell states that may reflect AD vulnerability independent of known risk factors, and thus warrant future mechanistic investigation. As such, we believe that perturbation experiments will be essential to disentangle whether these transcriptional signatures represent protective adaptations, pathogenic drivers, or reactive epiphenomena. These perturbation experiments, when coupled with our subcluster-level findings, open up avenues for more targeted therapeutic studies. Lastly, at present there remains a paucity of high-powered cohort data to examine case-control single cell status of AD ^56^, meaning future directions include expanding the current resources in sample size to further explore and resolve heterogeneity.

Overall, our work underscores the molecular complexity and heterogeneity of Alzheimer’s disease at single-cell resolution, revealing both convergent and cell-type–specific transcriptional programs of clinical AD and AD neuropathology that link genetic risk, demographic factors, and disease state. The discovery of genes independently supported by AD-associated SNPs and brain eQTLs provides a prioritized set of candidates for mechanistic follow-up and potential therapeutic targeting. Importantly, our results suggest that the molecular underpinnings of clinical and pathological AD are not wholly overlapping—reflecting either broader, pan-dementia processes or coupled but biologically distinct pathways within Alzheimer’s disease itself. By integrating these insights across cell types and disease definitions, this study offers a framework and resource to guide targeted interventions and refine our understanding of the cellular progression of AD.

## Methods

### Data source and cohort selection

We analyzed previously published single-nucleus RNA sequencing (snRNA-seq) data generated by Mathys et al. (2019), derived from post-mortem dorsolateral prefrontal cortex (PFC) tissue collected as part of the Religious Orders Study and Rush Memory and Aging Project (ROSMAP)^14^. We chose to use the prefrontal cortex for its important role in cognitive control and executive function. The original dataset included 48 individuals (26 with no clinical Alzheimer’s diagnosis, and 22 with clinical Alzheimer’s diagnosis) ^14^, balanced by age, sex, and educational attainment. Detailed clinical, cognitive, and neuropathological assessments were obtained longitudinally and at autopsy, as previously described ^7^. When classified based on CERAD pathology, there were 24 individuals with AD-like pathology and 24 individuals with non-AD pathology (Supplemental Fig. 3a). All participants provided informed consent, and protocols were approved by the Institutional Review Board of Rush University Medical Center.

SNRNA-seq libraries were prepared using the 10x Genomics Chromium Single Cell 3’ platform and sequenced using the NextSeq 500/550 system. Raw sequencing reads were aligned to the human reference genome (GRCh38.p5) using CellRanger (v2.0.0), including pre-mRNA counts to capture unspliced nuclear transcripts. After library-specific pre-processing, individual gene-count matrices were aggregated with read-depth normalization across samples. To assess potential technical confounding, we compared PMI between AD and control individuals and found no significant difference (FDR-adjusted p = 0.411), indicating comparable tissue preservation quality across diagnostic groups.

For our analyses, we obtained the processed gene-count matrix from Synapse (Synapse ID: syn18485175), consisting of an initial set of 17,926 genes profiled across 75,060 nuclei, as defined by Mathys et al. We applied additional quality control filters to exclude cells with low gene detection or aberrant mitochondrial read fractions, resulting in a final dataset comprising 17,926 genes and 70,634 high-quality nuclei. Cell-level metadata, including demographic variables and clinical annotations (e.g., age at first AD diagnosis, sex, APOE genotype), were integrated for each nucleus using unique barcode identifiers.

### Clinical and Neuropathological Definitions of Alzheimer’s Disease

We used two definitions of Alzheimer’s disease throughout our study, *clinical* Alzheimer’s diagnosis and *neuropathological* Alzheimer’s diagnosis. For our clinical diagnosis, we used the clinical diagnosis of cognitive status (DCFDX) rendered at the patient’s last visit via uniform, structured, clinical evaluation including a battery of 19 cognitive tests scored by computer, clinical judgement by a neuropsychologist, and diagnostic classification by a clinician. Using DCFDX, each patient is scored 1-6, with values of 4 or 5 corresponding to Alzheimer’s Dementia and a value of 6 corresponding to other primary causes of dementia, no clinical evidence of Alzheimer’s dementia. In our study, we used only scores of 4 or 5 to define clinical Alzheimer’s disease, treating other cases of “other primary cause of dementia, no clinical evidence of Alzheimer’s dementia” ^18^ as controls. For neuropathological based Alzheimer’s diagnosis, we used CERAD score which is a semiquantitative measure of neuritic plaques. For neuropathological Alzheimer’s, we required CERAD scores of 1 and 2, corresponding to frequent neuritic plaques (definite AD) in one or more neocortical regions and AD required moderate (probable AD), respectively ^57^.

### Cell type annotations in the Mathys and Lau et al data

For the Mathys et al. (2019) dataset, we used the available Broad cell type annotations (e.g., microglia, astrocytes, excitatory neurons) that were assigned using canonical marker gene expression profiles by Mathys et al. (2019). To further resolve cellular heterogeneity, we utilized subcluster labels from the Mathys et al. (2019) dataset.

For the Lau et al. (2020) validation dataset, we started with the single-nucleus gene expression matrices which we preprocessed and clustered using the Scanpy Python package. The expression matrix, containing 172,514 nuclei and 17,110 genes, was previously log-normalized. We constructed an AnnData object using these data without further normalization or log transformation. To reduce dimensionality while preserving biologically informative variability, we selected the top 5,000 highly variable genes (HVGs) using the ‘seurat’ flavor, which prioritizes genes based on normalized dispersion and mean expression. Total counts per nucleus (approximated by the sum of inverse-log-transformed expression values) and the proportion of mitochondrial gene expression per nucleus were regressed out using Scanpy’s pp.regress_out function to control for potential technical variation related to library size and cell quality, followed by scaling each gene to unit variance and zero mean. Principal component analysis (PCA) was performed on the HVGs, and the top 50 principal components were used to construct a k-nearest neighbors (kNN) graph with k = 30 neighbors. A two-dimensional t-distributed stochastic neighbor embedding (t-SNE) was computed using the top 10 PCs to visualize the manifold structure of the cells. Cells were clustered using the Louvain algorithm with a resolution parameter of 1.0, resulting in unsupervised cell clusters. For each cluster, we identified differentially expressed genes using a variance-adjusted t-test implemented in Scanpy’s tl.rank_genes_groups, comparing each cluster to all remaining cells. We selected the top 500 marker genes per cluster to perform statistical enrichment analysis against curated cell-type marker gene sets. Specifically, we evaluated the overlap between each cluster’s top marker genes and cell-type-specific marker lists for Astrocytes, Oligodendrocytes, Oligodendrocyte progenitor cells, Microglia, Excitatory neurons, and Inhibitory neurons. These marker lists were assembled from previously published high-confidence marker genes reported in Ianevski et al. (2022) and Lake et al. (2018) ^58,59^, and further refined by including canonical markers widely used in the Alzheimer’s disease single-nucleus RNA-seq literature (Mathys et al., 2019; Lau et al., 2020). Cluster assignments were further confirmed by systematically evaluating marker gene expression across clusters using dot plots and t-SNE overlays. Overlap was evaluated using a hypergeometric test (one-sided Fisher’s exact test), with enrichment p-values calculated per cell type and adjusted across all tested cell types within each cluster. The cell type with the most significant enrichment p-value was assigned as the label for the corresponding cluster. We compared our annotations to those reported in Lau et al. (2020) and found 90% concordance in annotations. To assess annotation robustness, we compared our cluster-derived cell-type labels to independent assignments obtained using a supervised marker-based approach using the marker gene annotations published by Lau et al. (2020). In the Lau-based approach, each cluster’s mean expression profile was scored against an external marker gene list, and cell types were assigned to clusters based on maximal cumulative marker expression. We observed high concordance between the two annotation strategies, with 155,278 out of 172,514 nuclei (90.0%) showing consistent labeling.

### TriSCOPE Predictive Modelling

We trained separate classifiers for each major cell type using normalized gene expression and evaluated their ability to predict Alzheimer’s status based on both Alzheimer’s diagnosis and CERAD neuropathological scoring. We trained on a variety of data modalities, including gene expression data and cell-level metadata variables: sex, education, age at death, and APOE genotype. We also tested various combinations of these data, including a combined model (gene expression data and all metadata variables), a gene expression data-only model, a metadata model (sex, education, age at death, and APOE genotype), and a gene expression and APOE genotype model. We also predicted on two sets of target variables, both Alzheimer’s diagnosis and CERAD pathology, using the National Institute on Aging’s guidelines for pathology-based diagnosis to group samples with CERAD scores of 1 and 2 as AD and scores of 3 and 4 as controls ^60^. We took strict steps to ensure no data leakage in any of our predictive models, this including grouped train/test splits by donor in all models. We ensured that all train-test splits were done sample wise, ensuring that all cells from a given sample were in either the training or testing data, but were not included in both. Every predictive model was run across 5 different random seeds, with the train-test splits being determined by the random seed. All presented findings were calculated using data from across these 5 splits to ensure robustness.

We developed a supervised machine learning framework to predict Alzheimer’s disease (AD) status at the single-cell level using the previously mentioned data modalities. All analyses were performed in Python (v3.9) utilizing the FLAML AutoML library (v1.2.0), scikit-learn (v1.3), and custom scripts. Single-nucleus RNA-sequencing data were processed to generate a cell-by-gene expression matrix, which was normalized and log-transformed prior to analysis. Metadata, including APOE genotype and clinical annotations, were integrated at the single-cell level using matching unique cell barcodes (TAGs). Post-mortem interval (PMI) was assessed as a potential confounder in both the Mathys et al. and Lau et al. datasets using non-parametric tests and violin plots (Supplementary Fig. 8). In both cohorts, PMI did not differ significantly between AD and control groups (FDR-adjusted *p* = 0.388 and *p* = 0.0949, respectively). Because PMI was constant across all cells from a given donor, and not associated with AD status, it was excluded from the predictive modeling framework to avoid overfitting and pseudoreplication. APOE genotype was encoded as a one-hot categorical variable. To identify individuals with AD, we defined case status based on the presence of a documented clinical diagnosis of Alzheimer’s disease prior to death, using metadata fields reflecting age at first AD diagnosis and related clinical variables. We performed a stratified train-test split at the donor level (80% train, 20% test) to prevent data leakage and to ensure that cells from the same individual did not appear in both sets. Stratification was performed jointly by AD status and sex to preserve class balance. Within only the training set data, we applied recursive feature elimination with cross-validation (RFECV) using a logistic regression estimator to select an optimal subset of informative genes, thus ensuring no data leakage. Genes with low mean expression or high sparsity were excluded prior to feature selection. After finalizing the gene set, expression data were merged with metadata features for each cell, and non-informative metadata variables were excluded to ensure model stability. The final feature set included the top-ranked genes along with APOE genotype encodings. We trained a gradient boosting classifier (LightGBM) using FLAML, adjusting the time budget based on the number of variables per training run. Model optimization was guided by log-loss and cross-validated using grouped folds defined by donor identity to further mitigate batch and individual-level confounding. To account for class imbalance, inverse-frequency sample weights were incorporated during training. Prediction probabilities on training and test cells were computed, and an optimal decision threshold was determined by maximizing the F1 score on the training set based on the precision-recall curve. Evaluation metrics included area under the receiver operating characteristic curve (ROC AUC), average precision, accuracy, recall, precision, F1 score, and Matthews correlation coefficient. To evaluate feature importance and robustness, we additionally performed incremental retraining using the top 1 to 25 features, assessing test performance at each step. Prediction probabilities, classification outcomes, and corresponding metrics were saved for downstream interpretability analyses, including subcluster-specific misclassification and differential expression assessments. External validation was performed using an independent snRNA-seq dataset comprising 12 AD patients and 9 controls (totaling 169,496 nuclei across microglia, oligodendrocytes, excitatory neurons, inhibitory neurons, and astrocytes). Validation analysis included two steps the first was fitting the predictive models trained on the Mathys et al dataset (to study generalizability of predictive modeling across cohorts) and the second part including rerruning our entire pipeline inorder to indepndelty discover which features were most important in the Lau et al dataset.

Validation analyses replicated the full predictive modeling workflow, including cell-type assignment, gene expression normalization, feature selection, and classifier training and evaluation. All performance metrics were calculated at the single-cell level, with train/test splits stratified and grouped by donor to avoid any cell from a given donor appearing in both sets.

Cell-level metrics were reported because the biological question centers on detecting disease-associated states of individual cells. Donor-level aggregation would obscure within-donor heterogeneity, which is the central focus of our study.

### Pathway analysis of predictive and differentially expressed gene features

To interpret the biological relevance of genes predictive of Alzheimer’s disease (AD) status, we performed pathway enrichment analysis using gene rankings derived from our cell type–specific LightGBM classifiers. For each major cell type (Astrocytes, Excitatory Neurons, Inhibitory Neurons, Microglia, Oligodendrocytes, and OPCs), we trained five independent models on randomized train/test splits. Feature importance scores from each split were extracted and normalized by the total non-zero importance values. For each gene, we computed the mean normalized importance across splits and retained genes with non-zero importance in at least two out of five splits. Non-gene features (e.g., demographic covariates or APOE encodings) were excluded using name-based filtering. We ranked predictive genes by their average normalized importance (using all genes with a non-zero feature importance in at least 2 splits) and for DEGs we performed gene set enrichment analysis (GSEA) using the prerank function in the GSEApy Python package (v1.1.0). Enrichment was conducted against the GO:Biological_Process_2023 gene set library from g:Profiler ^61^, using only the set of genes expressed in the filtered expression matrix as the background. Default parameters were used (minimum term size = 5, maximum = 1000), and a fixed random seed ensured reproducibility. The resulting pathways were filtered based on false discovery rate (FDR) < 0.05, and the top terms per cell type were visualized as barplots of –log₁₀(FDR). This same process was conducted for the differential expressed genes where applicable.

### SHAP-Based Model Interpretation Stratified by APOE Genotype

To identify the features most predictive of Alzheimer’s disease (AD) classification across distinct genetic risk backgrounds, we applied SHAP (SHapley Additive explanations) analysis to an integrated model trained on both gene expression and donor-level demographic features. This approach enabled us to dissect model behavior at single-cell resolution while accounting for individual-level covariates. For each major cell type (Astrocytes, Excitatory Neurons, Inhibitory Neurons, Microglia, Oligodendrocytes, and OPCs), we analyzed the output of five independently trained models, each derived from a stratified train-test split using a LightGBM classifier implemented via FLAML. Models were trained using both transcriptomic features (n = 17,926 genes) and demographic metadata (age at death, sex, years of education, and one-hot encoded APOE genotype) as previously described.

Unlike standard model interpretation methods that report global feature importance values from models trained solely on gene expression, our SHAP-based approach leverages a game-theoretic framework to compute the marginal contribution of each feature to the prediction for each individual cell. This allows for fine-grained, interpretable attribution of prediction decisions, enabling biologically meaningful comparisons across subgroups. To interrogate potential

APOE-specific transcriptional programs, we stratified all cells into three mutually exclusive genotype groups based on their donor’s APOE genotype: non-carriers of ε4 (e.g., ε2/3, ε3/3), heterozygous ε4 carriers (ε3/4), and homozygous ε4/ε4 carriers. For each group, we computed SHAP values on the subset of cells belonging to that group using the full model trained on all features. SHAP values were computed using TreeExplainer on the LightGBM booster model (.booster_) and then aggregated in two ways: Cell-Level Gene SHAP Scores: For gene expression features, we computed the absolute SHAP value for each gene in each cell and averaged across all cells in the group. This highlights the genes that most consistently influenced AD predictions within each genotype-defined subgroup. Donor-Level Metadata SHAP Scores: For demographic covariates (age, sex, education, APOE one-hot features), we computed the absolute SHAP values per cell and aggregated these by donor, then averaged across donors in each group. This strategy controls for donor-level sampling imbalance and isolates the contribution of metadata in shaping the model’s behavior.

Features were only retained for visualization and downstream analysis if they had non-zero SHAP variance across the five model splits and positive mean SHAP values, ensuring robustness and biological interpretability. Bar plots of the top metadata drivers (per donor) and gene drivers (per cell) were generated separately for each APOE group and each cell type. To emphasize highly predictive features, all bar plots display the square root of the mean absolute SHAP value across splits. This SHAP analysis extends beyond classical feature importance analysis by conditioning on APOE genotype and leveraging the full multimodal model rather than gene expression alone. It reveals genotype-specific predictive features that may not appear when aggregating across the full dataset, thus providing a mechanistically interpretable and genetically stratified view of the transcriptional programs driving AD predictions across distinct brain cell types.

### Differential expression analysis

We performed cell–type–specific differential expression (DE) analyses to identify genes associated with Alzheimer’s disease (AD) status across major brain cell types, including microglia, astrocytes, inhibitory neurons, excitatory neurons, oligodendrocytes, and oligodendrocyte precursor cells. For each cell type, cells were subset using broad cell type annotations. Gene expression matrices were first filtered to exclude genes with zero variance and genes expressed at low levels (counts per million [CPM] > 1 in fewer than 10 cells). For these analyses, raw UMI count matrices were used directly, with a log-transformed library size offset included in the regression to account for differences in sequencing depth.

We initially attempted to model gene-level expression using a Poisson mixed-effects model with a random intercept for donor (projid), aiming to account for inter-individual variability. We attempted Poisson/NB mixed-effects models with donor random intercepts, but saw frequent numerical instability and non-convergence. Consequently, we adopted a Poisson mixed effects model that explicitly controls for potential confounders while omitting the random effect term. For each gene, we fitted a zero-inflated Poisson model using the glmmTMB package (v1.1.7), including AD status as the primary variable of interest, and accounting for covariates for post-mortem interval (PMI), age at death, sex, and a library size offset (log total UMI counts per cell). We also checked our DEGs with a model adjusting for PMI, age, sex, APOE genotype, sequencing batch, and library offset size and found our results to be concordant. We report findings from the more conservative specification without APOE genotype and sequencing batch, which yielded a smaller set of DEGs. Cells with missing values for any covariates were excluded on a gene-wise basis. Estimated coefficients for AD status were extracted, and p-values were adjusted for multiple testing using the Benjamini-Hochberg procedure. Genes were considered differentially expressed if the FDR-adjusted p-value was less than 0.05 and the absolute log2 fold change exceeded 0.25. All analyses were performed in R (v4.2.2) using the glmmTMB, lme4, and dplyr packages.

To validate our single-cell differential expression results against a bulk RNA-seq standard, we generated pseudobulk profiles using Scanpy (v1.9.3). Within each broad cell type, raw UMI counts were summed across all cells from the same donor to create donor-level count matrices, retaining only donors with at least one cell in the given cell type. Gene expression was then filtered to retain genes with counts per million (CPM) ≥ 1 in at least 10% of donors (minimum of three donors). For each cell type, we fitted a linear model using the limma-voom pipeline (edgeR v3.40.2, limma v3.54.2), with Alzheimer’s disease (AD) status as the primary variable of interest and adjusting for the same covariates used in the single-cell models. Continuous covariates were mean-centered and scaled prior to model fitting. In parallel, we generated a “bulk-like” dataset by summing pseudobulk counts across all major cell types for donors with matched profiles across cell types, followed by the same limma-voom analysis. For both the per–cell-type and bulk-like analyses, moderated t-statistics for AD status were extracted to assess effect directionality and magnitude. We observed that the direction of log-fold changes in pseudobulk analyses generally matched those from our single-cell–level models, supporting the robustness of our single-cell findings to a bulk RNA-seq framework. Directionality agreement between single-cell and bulk differential expression results was 73.81% for astrocytes, 86.85% for excitatory neurons, 75.40% for inhibitory neurons, 57.73% for microglia, 76.86% for oligodendrocytes, and 78.72% for OPCs.

### GWAS enrichment and eQTL Analysis

To contextualize cell-type-specific transcriptional predictors within the known genetic risk architecture of Alzheimer’s disease (AD), we systematically evaluated their overlap with genome-wide association study (GWAS) loci and brain-specific expression quantitative trait loci (eQTLs). Significant AD-associated single-nucleotide polymorphisms (SNPs) were curated from the NHGRI-EBI GWAS Catalog (accessed via the term MONDO_0004975, “Alzheimer’s disease”). SNPs were mapped to associated genes using the mappedGenes field in the catalog.

For each major brain cell type, we compiled lists of predictive genes identified through our supervised machine learning classifiers. To ensure robustness, genes were retained only if they exhibited non-zero feature importance in at least two out of five cross-validation splits. Gene symbols were standardized to uppercase to ensure consistency in matching. As a background set for all enrichment analyses, we used the full set of 17,926 genes expressed in our single-nucleus RNA-seq dataset (Synapse accession: syn18485175), derived from the normalized expression matrix. This strategy controls for expression-dependent biases and provides a consistent denominator across analyses. To assess enrichment of GWAS signals, we performed one-sided Fisher’s exact tests (alternative = “greater”) comparing predictive genes with GWAS-mapped genes, relative to the background gene set, for each cell type. Overlapping genes, odds ratios, and p-values were recorded and visualized in Supplementary Tables. The full analysis was repeated analogously using predictors from classifiers trained to predict neuritic plaque burden using CERAD scores (pathology-based AD model). As with the clinical AD model, CERAD predictors were retained if non-zero feature importance was observed in at least two of five splits. To evaluate whether predictive genes were likely under genetic regulatory control, we assessed their overlap with cis-eQTLs identified in human brain tissue. We downloaded version 10 eQTL data from the Genotype-Tissue Expression (GTEx) project for three AD-relevant tissues: Brain – Cortex, Brain – Anterior cingulate cortex (BA24), and Brain – Frontal Cortex (BA9). We extracted Ensembl gene IDs with significant eQTLs (FDR < 0.05) and mapped these to gene symbols using a reference gene annotation file. Predictive genes were matched to eQTL genes based on shared gene symbols, and overlaps were identified per cell type. As with GWAS analysis, the background for Fisher’s exact tests was the full RNA-seq-expressed gene set (17,926 genes). To visualize eQTL support across cell types and predictive genes, we constructed a binary heatmap of the top predictive genes per cell type, indicating whether each gene had eQTL support in the three GTEx brain tissue. GWAS integration was performed by direct matching of exact genome-wide significant SNP rsIDs to model-nominated genes and brain tissue eQTL targets. Enrichment statistics (Fisher’s test p-values and odds ratios) were calculated for each cell type and for both clinical and CERAD-based models, and displayed alongside overlapping genes in Supplementary Fig. 10a–b.

### Pseudotime trajectory inference

For each cell subcluster containing at least 50 cells and representation from both Alzheimer’s disease (AD) and control individuals, we performed a pseudotime trajectory analysis to model a continuous transcriptional transition associated with disease progression. The analysis was restricted to a feature space defined by genes significantly differentially expressed (Benjamini-Hochberg adjusted *p* < 0.05) in the corresponding parent cell type, focusing on disease-relevant transcriptional signals. Gene expression data for each subcluster were normalized to 10,000 counts per cell, log-transformed (log(x + 1)), and processed using a standard Scanpy workflow. This included principal component analysis (PCA; up to 50 components), construction of a neighborhood graph (k = 10), and computation of a diffusion map. To define a biologically meaningful trajectory origin, we selected a root cell from individuals with no AD neuropathology (CERAD score = 4) ^62^ and no cognitive impairment (DCFDX last visit = 1) ^57^ within that pathological stage, defined as previously reported. This approach aimed to capture a putative early transitional state poised toward disease progression. To assess the stability of trajectory inference, the entire analysis pipeline, including the random sampling of a single root cell from the candidate pool, was repeated seven times using distinct random seeds — all calculated values were computed across these 7 runs to ensure robustness. For each iteration, diffusion pseudotime (DPT) was computed, and its association with AD diagnosis was quantified using two metrics: the area under the receiver operating characteristic curve (AUC-ROC) and a logistic regression model providing an odds ratio (OR) and corresponding *p*-value. Aggregated subcluster-level results included the mean AUC and mean log-OR, with 95% confidence intervals derived from standard errors across the seven runs. The *p*-values from each iteration were combined using Fisher’s method, and false discovery rate (FDR) correction was applied across subclusters using the Benjamini-Hochberg procedure. Finally, top trajectory-informing genes were identified by ranking genes based on the absolute loadings of the principal component which exhibited the highest positive correlation with pseudotime progression, ensuring alignment with the direction of disease-associated transcriptional change.

### Pathway activity scoring and correlation with pseudotime

We assessed whether transcriptional progression within each subcluster was associated with Alzheimer’s disease–related processes by testing a curated set of five Gene Ontology (GO) terms: amyloid precursor protein (APP) catabolism, amyloid-β formation, negative regulation of amyloid-β clearance, neuroinflammatory response, and positive regulation of microglial activation. GO annotations for each term were downloaded from QuickGO, with gene lists manually curated to remove duplicates and ensure consistent gene symbols. Expression data were normalized to 10,000 counts per cell and log-transformed using Scanpy’s normalize_total and log1p functions. Pathway activity scores were computed with Scanpy’s score_genes, which calculates the mean expression of the genes in a pathway relative to a background set of expression-matched genes (default of 25 bins) drawn from the remaining genes in the dataset.

This approach controls for baseline expression differences between genes. Within each subcluster, we computed the Spearman rank correlation between diffusion pseudotime values and pathway activity scores for the genes in each GO pathway module. False discovery rate (FDR) adjustment of p-values was performed using the Benjamini–Hochberg method across all subcluster–pathway pairs. Correlation coefficients and adjusted q-values were used to identify pathways whose activity was positively or negatively associated with pseudotime, and results were visualized as a pathway-by-subcluster heatmap.

### Validation Cohort

We validated our gene expression–based classifiers using an independent single-nucleus RNA-seq dataset derived from Lau et al., Proc Natl Acad Sci USA 2020 (PMID: 32989152), publicly available via NCBI GEO under accession number GSE157827. This dataset comprises gene-by-cell count matrices and accompanying metadata from 12 Alzheimer’s disease patients and 9 non-AD controls, generated from post-mortem prefrontal cortex tissue using standard single-nucleus RNA-seq pipelines as previously described by Lau et al, and containing Astrocytes, Excitatory Neurons, Inhibitory Neurons, Microglia, and Oligodendrocytes. We downloaded the processed expression matrix and metadata as released. We relied on the preexisting Lau et al marker genes to annotate our cells, however we performed independent cell-type annotation (as described above), to ensure consistency with our primary cohort and found strong concordance (90%). For our validation models, we analyzed a final set of 17,110 overlapping genes shared with our primary dataset. We firstly used the incremental classifiers fit on the Mathys et al dataset, froze these classifiers, and performed one-shot classification on the Lau et al dataset to examine predictive generalizability of the 1) Genes expression only classifier and 2) APOE Genotype + Gene expression classifier. For each cell type, classifiers trained on the full feature set and on top*-*k feature subsets (1–25) were evaluated on the Lau dataset, and the top classifier was reported. We then replicated our full predictive modeling workflow, including feature selection, cross-validation, and classifier training, across the 5 major brain cell types to independently assess feature stability. Consistent with our primary cohort, we observed strong predictive performance and robust convergence of top gene-level predictors, reinforcing the generalizability of our integrative framework.

### Quality-control validation on Murphy et al Dataset

We also performed a reanalysis of Mathys et al.’s work based on the cell types as defined by Murphy et al., after processing the data using scFlow ^63^. Taking this dataset of 50,381 “high-quality” nuclei taken from all 6 major cell types based on the filtering procedure previously described ^64^. During predictive modeling we dropped 941 nuclei that were missing post-mortem interval values. We found our results to be robust to this higher standard quality-control method, further validating our findings. Furthermore, we ensured that during our differential expression analysis we performed our analysis using poisson GLLMs, as done by Mathys et al, and prescribed it as a best practice, and equivalent to pseudobulk DE methods, for datasets with more than 500 cells ^7,12,64^. We found are pan-cell type predictors of ARL17B, LINGO1, RASGEF1B, and SLC26A3 to be predictive across cell types, and found cell type specificity for PLXDC2 in Microglia, GRID2 in Oligodendrocytes, and GFAP in Astrocytes. We additionally found predictive performance to be likewise similar.

## Supporting information

Supplementary Results

## Data Availability

All data produced in the present work are contained in the manuscript.
All primary data are available upon reasonable request.

https://adknowledgeportal.synapse.org/Explore/Studies/DetailsPage/StudyDetails?Study=syn3219045

## Data Availability

The snRNA-seq dataset supporting this study is publicly available from the Rush Alzheimer’s Disease Center (RADC) Research Resource Sharing Hub (https://www.radc.rush.edu/docs/omics.htm) or on Synapse (https://www.synapse.org/#!Synapse:syn18485175) under controlled access (DOI: 10.7303/syn18485175). ROSMAP metadata is accessible via Synapse (https://www.synapse.org/#!Synapse:syn3157322). Access requires a data use agreement to protect participant privacy.

The snRNA-seq dataset with further quality control measures applied to the Mathys et al dataset supporting this study is publicly available on Synapse (https://doi.org/10.7303/syn51758062.1). Data is available for general research use according to the following requirements for data access and data attribution (https://adknowledgeportal.org/DataAccess/Instructions).

The validation single-nucleus RNA-seq dataset supporting this study is publicly available from the NCBI Gene Expression Omnibus under accession number GSE157827 (https://www.ncbi.nlm.nih.gov/geo/query/acc.cgi?acc=GSE157827). This dataset comprises prefrontal cortex samples from 12 Alzheimer’s disease patients and 9 non-AD controls, and was originally published in Lau et al., Proc Natl Acad Sci USA 2020 (PMID: 32989152). Access to raw data requires compliance with relevant data use agreements.

The brain frontal, prefrontal, and cortex tissue eQTL data used for the analyses described in this manuscript were obtained from the GTEx Portal on 07/26/25.

## Code Availability

https://github.com/AdiVM/TriSCOPE_RNASeq_Alzheimers_Analysis

The code used in this study has been deposited in the private GitHub repository above. To facilitate peer review, we have included a zip file with the entire repository’s codebase and documentation. All code will be made publicly available in the GitHub repository after peer review.

## Supplementary Table Captions

**Supplemental Table 1:** Feature importance values per cell type across the five experimental splits for a model that used demographics, APOE genotype, and gene expression data to predict clinical AD.

**Supplemental Table 2:** Feature importance values per cell type across the five experimental splits for a model that used gene expression data to predict clinical AD.

**Supplemental Table 3:** Feature importance values per cell type across the five experimental splits for a model that used demographics, APOE genotype, and gene expression data to predict AD neuropathology.

**Supplemental Table 4:** AUC summary again on a per cell type level basis, 4 for clinical AD, 4 for AD neuropathology, 2 for the validation dataset.

**Supplemental Table 5:** Overlap between the GWAS catalog mapped genes and predictively expressed clinical AD genes on a per cell type basis.

**Supplemental Table 6:** Overlap between the GWAS catalog mapped genes and predictively expressed in AD neuropathology genes on a per cell type basis.

**Supplemental Table 7:** Fisher’s exact test results for enrichment of predictive AD genes that did overlap with GWAS loci. Values are shown per cell type, reportingthe significance of overlap across independent predictive and differential expression analyses.

**Supplemental Table 8:** Differential expression results for clinical AD versus controls across all major brain cell types. Includes log₂ fold changes and FDR-adjusted p-values on a per cell type basis.

**Supplemental Table 9:** Differential expression results for AD neuropathology across all major brain cell types. Includes log₂ fold changes and FDR-adjusted p-values.

**Supplemental Table 10:** Overlap of predictive AD genes between the Mathys and Lau datasets, reported on a per-cell-type basis. Genes that emerged as predictive in both datasets are highlighted.

**Supplemental Table 11:** Fisher’s exact test results for validation analyses, assessing enrichment of overlapping predictive and DE genes between the Mathys and Lau datasets.

**Supplemental Table 12:** Differential expression results for the validation dataset (Lau et al.), reported per cell type. Includes log₂ fold changes and FDR-adjusted p-values.

**Supplemental Table 13:** Feature importance values per subcluster across the five experimental splits for models trained on gene expression to predict clinical AD. Highlights fine-grained subpopulation-level predictors.

**Supplemental Table 14:** Summary statistics from pseudotime analyses, including trajectory significance, correlation with pathway activity, and subcluster-level pseudotime associations with AD outcomes.

**Supplemental Table 15:** Full donor-level metadata summary across both cohorts (Mathys and Lau), including age at death, sex, education, APOE genotype, clinical diagnosis, CERAD neuropathology score, and post-mortem interval.

## Supplemental Tables

- **1. Supplemental Table 1: AD_CombinedModel_Feature_Importances_CellLevel.xlsx**
- **2. Supplemental Table 2: AD_GenesModel_Feature_Importances_CellLevel.xlsx**
- **3. Supplemental Table 3: CERAD_GenesModel_Feature_Importances_CellLevel.xlsx**
- **4. Supplemental Table 4: AUC summary**
- **5. Supplemental Table 5: GWAS_AD_overlap_summary.xlsx**
- **6. Supplemental Table 6: GWAS_CERAD_overlap_summary.xlsx**
- **7. Supplemental Table 7: All_NonGWAS_Fisher_Tests.xlsx**
- **8. Supplemental Table 8: AD_Differential_Expression.xlsx**
- **9. Supplemental Table 9: CERAD_Differential_Expression.xlsx**
- **10. Supplemental Table 10: Validation_Overlap_Results.xlsx**
- **11. Supplemental Table 11: Validation_Fisher_Tests.xlsx**
- **12. Supplemental Table 12: Validation_Differential_Expression.xlsx**
- **13. Supplemental Table 13: AD_GenesModel_Feature_Importances_SubclusterLevel.xlsx**
- **14. Supplement Table 14: Pseudotime_summary_statistics.csv**
- **15. Supplemental Table 15: Full Donor Data Summary**

## Supplementary Figure Captions

**Supplementary Figure 1: Validation of predictive classifiers in an independent cohort**

**a,** Scatterplots comparing normalized predictive feature importance values for genes independently identified in both the Mathys et al. and Lau et al. datasets across five major brain cell types (Astrocytes, Excitatory Neurons, Inhibitory Neurons, Microglia, and Oligodendrocytes). Genes are plotted by their mean normalized importance in Mathys (x-axis) versus Lau (y-axis), with overlap counts indicated in each panel. **b,** Receiver operating characteristic (ROC) curves showing “one-shot” classification performance of classifiers trained on Mathys data, and tested on the independent Lau dataset. Results are shown for classifiers using gene expression only (blue) or gene expression plus APOE genotype (orange). **c,** Bar plots showing the top-ranked predictive features for classifiers independently trained de novo on the Lau dataset across the same five brain cell types. **d,** Volcano plots showing differentially expressed genes (DEGs) between Alzheimer’s disease and control cells across the Lau dataset, analyzed separately by cell type. Genes significant for differential expression only are shown in blue, predictive-only genes are in grey, and genes that were both predictive and differentially expressed are highlighted in red. **e,** Heatmap showing log₂ fold changes of the top 80 DEGs across cell types. Genes were selected by absolute fold change and statistical significance, and ranked across cell types to visualize cross-cell-type convergence and divergence of transcriptional alterations. Color intensity indicates log₂ fold change, with red = upregulation and blue = downregulation.

**Supplementary Figure 2: Differential expression analysis across combined clinical–pathological groups.**

**a,** Volcano plots showing differentially expressed genes (DEGs) across four clinical–pathological contrasts, performed separately for each of the five major brain cell types (Astrocytes, Excitatory Neurons, Inhibitory Neurons, Microglia, Oligodendrocytes, and Oligodendrocyte Progenitor Cells). The four contrasts tested were: (i) Clinical AD versus Clinical Control within pathology-negative individuals, (ii) Pathology AD versus Pathology Control within clinically non-demented individuals, (iii) Clinical AD versus Clinical Control within pathology-positive individuals, and (iv) Pathology AD versus Pathology Control within clinically diagnosed AD cases. Genes significant for differential expression (FDR-adjusted *P* < 0.05) are colored in red (upregulated) and blue (downregulated). **b,** Pathway enrichment analysis of DEGs identified in Microglia for the contrast of Pathology-positive Clinical AD versus Pathology-positive Clinical Controls. **c,** Pathway enrichment analysis of DEGs identified in Oligodendrocytes for the contrast of Pathology-positive Clinical AD versus Pathology-positive Clinical Controls.

**Supplementary Figure 3: Demographic, compositional, and clustering characteristics of Mathys and Lau datasets.**

**a,** Distribution of donors across clinical and neuropathological categories in the Mathys dataset, showing separation into four combined groups (Clinical Control/Pathology Control, Clinical AD/Pathology Control, Clinical Control/Pathology AD, and Clinical AD/Pathology AD). **b,** Cell type composition across both Mathys and Lau datasets, demonstrating broadly consistent representation of astrocytes, excitatory neurons, inhibitory neurons, microglia, and oligodendrocytes. **c,** Age-at-death distributions for AD cases and controls across both cohorts, indicating similar demographic ranges. **d,** UMAP projection of the validation (Lau) dataset annotated using marker genes from the original study, showing robust separation of major brain cell types. **e,** Validation dataset annotated with Mathys marker genes, demonstrating concordance in broad cell type assignment with panel D. **f,** Leiden clustering of the Lau dataset, capturing additional substructure within annotated cell types. **g,** UMAP projection of the Mathys dataset annotated with broad cell types. **h,** Subcluster-level annotation of the Mathys dataset, showing finer stratification within major cell classes. **i,** Leiden clustering of the Mathys dataset, highlighting resolution of transcriptionally distinct subpopulations. **j,** Post-mortem interval (PMI) distributions for clinical AD versus control donors in the Mathys dataset, showing no significant difference between groups. **k,** PMI distributions for clinical AD versus control donors in the Lau dataset, similarly showing no significant difference.

**Supplementary Figure 4: SHAP-based interpretation of predictive models for clinical AD status stratified by APOE genotype**

**a**, Bar plots showing the top gene-level predictors of clinical AD classification across major brain cell types identified by SHAP analysis. **b**, Heatmaps displaying per-gene SHAP ranks across APOE genotype groups (non-carriers, heterozygous ε4, homozygous ε4/ε4). **c**, Radar plots summarizing APOE-stratified SHAP values for selected top genes within each major cell type, showing shifts in predictive contribution between genotype-defined subgroups. **d**, Heatmap showing Spearman correlation between gene expression and predicted AD probability across cell types for a representative subset of key predictors. Positive correlations highlight cell-type–specific associations between expression level and model-predicted AD probability, whereas negative correlations suggest context-dependent or compensatory roles.

**Supplementary Figure 5: Linking cell-type–specific predictive features to AD GWAS and brain eQTL support.**

**a,** Heatmap of clinical AD predictors (identified in our modeling framework) that overlap with genome-wide significant AD GWAS loci (P < 5 × 10⁻⁸) and additionally exhibit support from brain tissue eQTL datasets. Predictors are shown in a cell-type–specific manner, and the eQTL evidence was derived from bulk brain tissue rather than cell-type–specific eQTL resources. **b,** Heatmap of AD neuropathology predictors overlapping with GWAS loci and brain eQTL support, analogous to panel A. **c,** Heatmaps showing –log₁₀ GWAS P-values for top neuropathology-associated predictors overlapping with GWAS loci, stratified by cell type. Up to 10 most significant genes are shown per cell type. Notably, APOE exhibits strong GWAS association in both astrocytes and microglia. **d,** Venn diagrams illustrating the overlap between clinical AD GWAS-supported predictors and neuropathology GWAS-supported predictors across cell types, highlighting convergent and distinct sets of genetically anchored disease-relevant features.

