## Supplementary Results for "Single-cell machine learning uncovers genetically anchored, cell-type specific programs of Alzheimer’s disease"

### Supplementary Information

Madduri et al

#### Supplementary Results

**Clinical AD Prediction**

Our AD classifiers revealed significant cell-level heterogeneity with Microglia (mean AUC: 0.653 ± 0.085), Inhibitory Neurons (mean AUC: 0.628 ± 0.074) and Oligodendrocytes (mean AUC: 0.652 ± 0.096) all demonstrating significant predictive performance using gene expression data alone relative to demographics (Supplemental Table 16, Fig. 2a). Classifiers trained on APOE genotype (without demographics) and gene expression data, exhibited significant predictive power in all cell types, and outperformed classifiers trained on Demographics and APOE genotype in Astrocytes, Oligodendrocytes, Inhibitory Neurons, and Microglia (Supplemental Table 16, Fig. 2a). To identify the molecular drivers underlying each model's performance, we next ranked feature importance scores across all cell types, both in our combined data modality models and gene expression-only models. Across our combined data modality models, age at death, education, APOE ε4/ε4 genotype, APOE ε4 homozygous genotype, sex, *ARL17B*, *HSPA1A*, *RASGEF1B*, and *LINGO1* were the most predictive features across cell types (Fig. 2c). These findings reinforce that while demographics contribute to disease prediction, they alone do not explain the transcriptional heterogeneity observed in AD on a cell-type basis.

Notably, several genes repeatedly ranked among the most important predictors across cell types, including *RASGEF1B*, *LINGO1*, and *ARL17B* (Fig. 2c, d). These genes remained top-ranked even after accounting for demographic and APOE genotype, suggesting that they reflect intrinsic molecular signals of disease. *RASGEF1B*, a member of the RAS signaling pathway, which has an established role in synaptic plasticity and neuronal dysfunction, emerged as a potential cross-cell-type effector of AD vulnerability (Stornetta and Zhu 2011; Shankar and Walsh 2009). *LINGO1* is a known regulator of axonal regeneration and myelination, and has been implicated Alzheimer's disease (Fernandez-Enright and Andrews 2016). *ARL17B*, which encodes Rapidly Accelerated Fibrosarcoma (RAF) GTPases, likewise demonstrated pan-cell-type importance consistent with links between the GTP-binding protein complex and amyloid precursor protein (APP) to disease progression[21,22,23].

While some features were shared across cell types, others were highly specific. Top predictors that were highlighted in only one cell type included (SUPPLEMENTARY TABLE XXX): In Excitatory Neurons, we uniquely found that *MTRNR2L1* and *MTRNR2L8* were top predictors (Fig. 2b,  Supplemental Table 1). *MTRNR2L1* (also known as HN1) encodes Humanin, a mitochondrial-derived peptide linked to Alzheimer's, Parkinson's, and other neurodegenerative disorders due to its neuroprotective effects, including inhibition of neuroinflammation [19].

In Inhibitory Neurons, we identified *AHI1* as a uniquely strong predictor. *AHi1* has been linked to impaired cognition and reduced synaptic plasticity in prenatally stressed offspring[24], and its importance here suggests a potential role in neuronal dysfunction in AD. *SLC26A3,* chloride/bicarbonate exchanger with emerging roles in neuronal physiology, also emerged as a top predictor in Inhibitory Neurons (Fig. 2b). Notably, we also observed an enrichment of Y chromosome–linked genes, including *USP9Y*, *NLGN4Y*, and *UTY*, suggesting a possible male-specific transcriptional program related to AD vulnerability in this population of neurons (Supplemental Table 1).

In Microglia, we identified a unique set of top-ranked predictive genes, suggesting that this cell type may follow a distinct disease-induced transcriptional trajectory. The strongest predictor was *IFI44L*, a gene previously linked to interferon signaling (Jin et al. 2022) and immune activation in the brain (Fig. 2b, Supplemental Table 1). Other highly ranked features included *SRGN*, *HIF1A*, and *APOE*, (Qian et al. 2024). *PLXDC2*, a gene associated with immune response in tumor microenvironments, also emerged as a strong predictor, highlighting a potential immune effector role in Microglial responses to AD (Supplemental Fig. SHAPa, Supplemental Table 1). Notably, *PTEN*, a gene previously shown to bind *PLXDC2* in macrophages to promote inflammatory activation and prominent in GWAS, was also predictive. Another top-ranked feature was *HSPA1A*, a molecular chaperone that has been linked to protein quality control and the inhibition of amyloid-β and neurofibrillary tangle formation[30]. Collectively, these predictors support a Microglia-specific molecular signature enriched for immune and stress-response regulators (Fig. 2b, Supplemental Table 1).

In Oligodendrocytes, top predictive features included *CRYAB*, *CLDN11*, and *APOD* all of which were better stronger predictors than APOE Genotype e3/e4 status (Supplemental Fig. 10; Fig. 2b; Supplemental Table 12). *CRYAB* encodes a molecular chaperone linked to stabilization of intermediate GFAP filaments and more generally linked to prevention of protein aggregation (Muraleva et al. 2019; Becerra-Hernández et al. 2022) possibly representing a response to protein aggregation and misfolding events in Alzheimer's (Rudenko et al. 2019). In OPCs, three top targets, *APOD, FTH1*, and *COL4A3* emerged (Fig. 2d; Supplemental Table 1; Supplemental Table 12). In Astrocytes, top predictive features included *ID2*, *MT2A*, *FTH1*, and *GFAP* (Fig. 2b, Supplemental Table 1). *ID2*, a gene encoding Inhibitor of DNA Binding 2, is known to be involved in inflammatory signalling, possibly linking Astrocytic inflammatory pathways in

Alzheimer's to previous research that showed under inflammatory conditions cytokines upregulate *ID2* (Fig. 2b) [34]. *GFAP* is a well-established astrocyte marker and has previously been used as an Alzheimer's biomarker (Kim et al. 2023). The identification of well-characterized AD-related genes among top features reinforces the biological validity of our modeling approach, and suggests that those high-ranking predictors with limited prior characterization, may also playing meaningful roles in AD (Fig. 2e).

#### DEG Analysis

To contextualize and further interpret features identified by predictive modeling, we performed differential expression analysis between clinical AD and control samples within each major cell type.

In Excitatory Neurons, two of our top predictors *LINGO1* and *RASGEF1B*, were both differentially expressed in Alzheimer's disease (AD) cases. LINGO1 showed significant upregulation ($log_2FC = 0.28$, FDR = 2.3E-209) and RASGEF1B was also marginally upregulated ($log_2FC = 0.26$, FDR = 1.6E-179) (Fig. 3c) (Supplemental Table 6). In addition, *MTRNR2L1* ($log_2FC = 1.39$, FDR = 2.45E-82) was significantly upregulated and *MTRNR2L8* was significantly downregulated ($log_2FC = –0.39$, FDR = 2.44E-17) (Supplemental Table 6). While differential expression analysis revealed over 1,081 genes (Fig. 3b) altered in AD within Excitatory Neurons, a much smaller subset of only 162 features (Fig. 4e) were prioritized (24 overlapping, Fig. 3a) by our predictive modeling analyses, highlighting a much more focused core of disease-relevant changes when using a multivariate machine learning approach as opposed to a univariate differential expression approach. In Inhibitory Neurons, 17 of our predictive features were significantly differentially expressed in Alzheimer's disease (AD) cases. Though we identified 252 DEGs, many of our highly predictive features — including *AHI1*,

*USP9Y*, *NLGN4Y*, and *UTY* — did not meet differential expression thresholds (FDR < 0.05) (Fig. 3c, Supplemental Table 6). An exception was SLC26A3, which was significantly upregulated (FDR = 1.38E-27, FDR = 0.40) (Supplemental Table 6)

In Microglia, several top predictive genes were also significantly differentially expressed between Alzheimer's disease (AD) and control samples. *IFI44L*, the highest-ranking predictor, was significantly downregulated ($log_2FC$ = –1.21, FDR = 0.0006), consistent with prior evidence implicating impaired type I interferon signaling, and reduced expression of interferon-stimulated genes like *IFI44L*, in individuals at elevated risk of AD progression (Song et al. 2022). SRGN, another strong immune-related predictor, was markedly downregulated ($log_2FC$ = –1.30, FDR = 6.78E–19) (Fig. 3c, Supplemental Table 6). *HSPA1A*, **a** chaperone gene associated with protein quality control, was also significantly reduced ($log_2FC$ = –0.57, FDR = 3.56E-05), alongside upregulation of APOE ($log_2FC$ = 0.36, FDR = 0.009). In contrast, PLXDC2 and PTEN—both of which showed predictive importance—were not significantly differentially expressed.

In Oligodendrocytes, *HSPA1A* — one of the top predictive features — was significantly downregulated in Alzheimer's disease (AD) cases, showing robust downregulation ($log_2FC$ = -0.55, FDR = 2.34E-35) (Fig. 3c, Supplemental Table 6). In total, we observed 92 predictive and differentially expressed genes, with 16% of predictive genes being DEGs. In contrast, other top-ranked predictors such as *APOD* were moderately differentially expressed, while *CRYAB* was not differentially expressed (Supplemental Table 6). These results highlight the potential of predictive modeling to uncover disease-relevant features that may not exhibit strong univariate shifts but could participate in multivariate, context-dependent expression patterns in AD. In Oligodendrocyte Progenitor cells (OPCs), two of the top predictive features, *FTH1* and *APOD*, were also significantly differentially expressed in Alzheimer's disease (AD) cases. *FTH1* was upregulated with a $log_2$ fold change ($log_2FC$) of 0.37 (FDR = 8.41E-06), and *APOD* similarly showed significant upregulation ($log_2FC$ = 0.27, FDR = 2.54E–08) (Fig. 3c, Supplemental Table 6). In Astrocytes, *MT2A* and *GFAP*, were significantly differentially expressed in Alzheimer's disease (AD) cases. *MT2A* was also strongly upregulated ($log_2FC$ = 0.57, FDR = 3.82E-40). *GFAP*, a known astrocytic marker, exhibited significant upregulation ($log_2FC$ = 0.45, FDR = 1.93E-11) (Fig. 3c, Supplemental Table 6).

Overall, we have identified robust transcriptional changes in a cell-type specific manner between Alzheimer's patients and controls, defined by clinical adjudication. Our prior analyses of predictive genes highlighted genes with varying overlap with DEGs (2.75-32% of predictive genes were DEGs in each cell type). Together, these findings demonstrate that while differential expression analysis captures many transcriptional changes in Alzheimer's disease (AD), our predictive modeling approach identifies a complementary but often non-overlapping set of biologically meaningful features. In most cases, these include genes that are subtly dysregulated or fall beneath conventional significance thresholds but may reflect important disease mechanisms—such as sex-specific vulnerability programs in Inhibitory Neurons or immune-related regulatory shifts in Microglia. These results reinforce the ability of predictive modeling to nominate both strongly altered and more nuanced transcriptional signals involved in AD neuropathology, providing additional insight into the cell-type–specific molecular architecture of disease.

#### Clinical and Pathology Predictors Diverge Across Cell Types

Considering the two key hallmarks of Alzheimer's, cognitive decline and amyloid, tau, and neurodegenerative (ATN) pathology, we suspected that gene expression and other biological features may explain neuropathology-based AD diagnosis differentially from clinicly-based AD diagnosis (Supplemental Fig. 3a). We tested this hypothesis by rerunning our analysis of the Mathys et al dataset[10], with outcomes and group labels defined by neuropathology (CERAD score 1-2) rather than clinician diagnosis. Specifically, we predicted moderate or frequent neuritic plaques based on CERAD score (i.e., CERAD: 1 or 2 as AD), a semiquantitative measure of neuritic plaque pathology. Our classifiers revealed that gene expression and APOE genotype strongly predict neuritic plaque pathology across cell types (Fig. 4a).

In Astrocytes (mean AUC: 0.691 ± 0.063), we found top predictors to include *RASGEF1B*, *MT1E*, and *GFAP* as well as *APOE*. We also observed predictive power in Excitatory Neurons (mean AUC: 0.739 ± 0.179), Inhibitory Neurons (mean AUC: 0.695 ± 0.196), and Microglia (mean AUC: 0.645 ± 0.188) (Fig. 4a). We found strong overlap between genes predictive of amyloid-plaque pathology and clinical diagnosis; however, their importance was rearranged. In Microglia, top predictors included *SAT1*, *PRKCA*, and *APOE*. *SAT1* (log2FC = -0.44, FDR p = 5.76E-08), a gene whose increased expression has previously been linked to immune cell infiltration in glioma, was downregulated in AD, complementing our previous finding of IFI44L downregulation [35] (Fig. 4b, Supplemental Table 7). *PRKCA*, a gene responsible for lipopolysaccharide-induced macrophage functions, including inflammation and defense, was also downregulated (Fig. 4c, Supplemental Table 2). *APOE* was significantly upregulated, consistent with its role in advanced Amyloid pathology (Supplemental Table 7).

Overall, we demonstrate complementary findings across our predictive and differential expression approaches for both clinical and neuropathology outcomes. First, despite cell-to-cell heterogeneity, *RASGEF1B* was observed as a top predictor of both CERAD pathology and clinical diagnosis in all cell types. Second, we observed strong concordance between genes predictive of pathology AD and clinical AD across cell types (Fisher's $q < 1.92e-20$) (Supplemental Table 5), but especially in Microglia with 244 overlapping predictors (OR = 106.7, 95% CI: 84.4-134.9) and in Oligodendrocytes 238 overlapping predictors (OR = 33.09, 95% CI: 27.2-40.27) (Fig. 4e, Supplemental Table 5). These results suggest in Microglia and Oligodendrocytes, molecular signals of amyloid pathology may be tightly coupled to clinical manifestation of disease (Leng and Edison 2021; Kedia and Simons 2025).

GWAS Integration:

We tested whether genome-wide significant AD single-nucleotide polymorphisms SNPs from the NHGRI-EBI GWAS Catalog exert functional effects on transcriptomic predictors of both clinical and pathological AD by identifying rsIDs that (i) map to model-nominated genes, and (ii) checking if those exact SNPs were eQTLs in human brain tissue.

Clinical AD predictors in Inhibitory Neurons, Microglia, Oligodendrocyte Progenitor Cells, and Oligodendrocytes were significantly enriched for genes containing genome-wide significant SNPs (Fisher's p range=1.78e-5 - 0.047). Our analyses revealed pan-cell-type genetic convergence, including LINGO1 (rs12902811) (-log10 p = 13.7) and ARL17B (rs2732703) (-log10 p = 8.22) (Fig. 3e, Supplemental Table 3). The gene in Oligodendrocytes with the strongest association with AD was *CLU* (rs11787077) (-log10p=32), and *CLU* gene expression levels have been implicated in cognitive decline in Alzheimer's (Foster et al. 2019) (Supplemental Table 3). Microglia showed 68 predictive overlapping GWAS genes (OR = 1.48,

Fisher's p = 0.047), including *APOE* (-log10p = 3.2E+02), *BIN1* (1.2E+02), *PICALM* (rs561655) (-log10p = 26), and *RASGEF1C* (rs113706587) (-log10 p = 15.7) (Fig.3e, Supplemental Table 3). ABCA1 was a top GWAS hit in Astrocytes (-log10p = 32). To further examine functional relevance, we assessed whether these predictive GWAS-associated genes were also regulated at the transcriptional level using the GTEx brain tissue eQTL databases (GTEx Consortium 2020). We found that across all cell types, 5 SNPs had both eQTL's in prefrontal cortex relevant tissue and were GWAS associated genes functionally predictive of Alzheimer's. Of these, *APOE* (rs769446), *CEP170* (rs71537331), *CTSB* (rs1065712), and *PICALM* (rs17745409) were predictors in Microglia, while *CLU* (rs4236673) was predictive in Oligodendrocytes and Excitatory Neurons. *CLU* and *PICALM* (rs4236673) were predictive in Oligodendrocyte Progenitor Cells.

We repeated this overlap analysis using predictors of AD neuropathology rather than clinical diagnosis, which revealed *APOE* as the top hit in both Astrocytes and Microglia (Supplemental Fig. 6c, Supplemental Table 4). We once again found the strongest overlap with GWAS hits of 107 genes in Oligodendrocytes (OR = 1.85, Fisher's p = 1.27E-07) (Supplemental Table 4). We then examined the overlap between clinical and pathological AD predictive genes that were also genome-wide significant in the GWAS catalog. In Excitatory Neurons, all pathology-associated GWAS hits also appeared among clinical AD GWAS hits and showed LINGO1 to be a shared GWAS hit in every cell type (Supplemental Fig. 6d). *CLU* was shown to be only present in the clinical AD GWAS set, suggesting a potential bias toward cognitive AD phenotypes. *APOE* overlapped between pathology- and clinical-AD GWAS hits in Microglia, but was only observed in the pathology-associated GWAS set in Astrocytes. Our CERAD predictors also showed highly significant overlap (Fisher's p = 1.36E-245) with brain tissue eQTL's and revealed similar sets of genes (Supplemental Fig. 10b). Our analysis thus reveals that there is heterogeneity among the functional effects of GWAS associated genes at the mRNA level, suggesting that GWAS-associated genes may exert differential, cell-type-specific effects, and may variably influence clinical or pathological manifestations of AD. We then took these GWAS associated pathological predictors and checked for brain tissue eQTLs, revealing 5 SNPs. These included *APOE* in Astrocytes, *APOE* and *PICALM* in Microglia, *CEP170* and *CEP63* in Inhibitory Neurons, and *ARFGEF3* in OPCs. By finding direct rsID overlap, we establish a functionally interpretable link between these previously GWAS associated and gene expression that is predictive of Alzheimer's disease. Furthermore, heterogeneity between Clinical and Pathological Alzheimer's predictors, may hint at differing stages or manifestations of disease severity.

To better understand these differences, we performed differential expression analysis between individuals who exhibited Alzheimer's pathology, but no clinical diagnosis, and those with Alzheimer's pathology and clinical diagnosis. Our analysis revealed an upregulation of *SRGN*, *HIF1A*, and *RASGEF1C* in Microglia, and subsequent pathway analysis revealed enrichment of pathways related to neuroinflammatory response, cytokine production and interleukin-6 regulation (Supplementary Fig. 2a, b). Similar analysis in Oligodendrocytes revealed upregulation of pathways related to cellular response to unfolded and topologically incorrect protein and pathways related to long-term memory (Supplemental Fig. 2c).

**Validation**

To confirm the robustness of these findings, we validated our cell-type-specific classifiers on an independent dataset comprising 12 clinical AD and 9 clinical control individuals, covering 169,496 nuclei across Microglia, Oligodendrocytes, Excitatory Neurons, Inhibitory Neurons, and Astrocytes (Supplementary Table 9). Using one-shot classifiers from our original dataset, and ensuring strictly no donor level data-leakage, we showed consistent predictive performance across cell types suggesting the generalizability of our models (Supplementary Fig. 1a). Our validation ethclassifiers demonstrated consistent performance across cell types, with gene-only one shot classifiers showing mean AUCs greater than 0.5 in Astrocytes ( AUC 0.67), Oligodendrocytes (gene-only classifier mean AUC 0.58), Inhibitory Neurons (gene-only classifier mean AUC 0.60), and Excitatory Neurons (gene-only classifier mean AUC 0.65) (Supplementary Fig. 1b). More significantly, upon retraining our models we observed strong concordance in top predictive genes, once again with cell type specificity. These included *NLGN4Y* in Inhibitory Neurons and *MTRNR2L8* in Excitatory Neurons (Supplementary Fig. 1c, Supplemental Table 9) . Similarly Microglia showed *PLXDC2* (log2FC = 0.22) and *APOE* (log2FC = 0.857) as top predictors and DEGs (Supplemental Fig. 1d), while Astrocytes showed *COL21A1* and *GFAP*. In Oligodendroctes we observed *CRYAB*, *FTH1*, and *APOD* to all be top predictors. We once again saw *RASGEF1B*, *ARL17B*, *LINGO1*, and *SLC26A3* as predictive across multiple cell types, reinforcing their robustness as pan-cell-type AD predictors (Supplemental Table 10, Supplemental Fig. 1b)

We observed strong predictor overlap between our original and validation datasets across all five major cell types (Fisher's Exact Test, Supplemental Table 9). In Inhibitory Neurons, *RASGEF1B*, *ARL17B*, *NLGN4Y, SLC26A3, and LINGO1* were top predictors, and differential expression analysis revealed all of these genes to be differentially expressed (FDR adjusted $p < 0.05$) (Supplemental Table 11). In Excitatory neurons top predictors were *PDE10A*, *RASGEF1B*, *NLGN4Y*, *SLC26A3*, *USP97*, *ARL17B*, *MTRNR2L8*, and *LINGO1*. Our validation analysis in Astrocytes showed top predictors as *SLC1A2*, *COL21A1*, *SLC26A3*, *RASGEF1B*, *GFAP*, and *HSPA1A*. In Microglia, *PLXDC2* and APOE once again emerged as cell-specific predictors (Supplemental Fig. 1c), with *APOE* (log2FC = 0.857) and *PLXDC2* (log2FC = 0.22) both significantly differentially expressed (FDR adjusted $p < 0.05$) (Supplemental Fig. 1c). Across the five cell types, we found strong overlap in predictive genes (FDR adjusted Fisher's $p < 0.05$ in all cell types) (Supplemental Fig. 1a, Supplemental Table 10), including 40 overlapping predictors in Astrocytes (OR = 30.28), 14 in Excitatory Neurons (OR = 560.06), 12 in Inhibitory Neurons (OR = 415.55), 122 in Microglia (OR = 40.39), and 106 overlapping in Oligodendrocytes (OR = 21.78) (Supplemental Fig. 1a, Supplemental Table 10).

#### Subcluster Profiles Reveal Transcriptional Gradients of AD Progression

After discovering genes both predictive and differentially expressed in clinically and neuropathologically-defined AD, we examined whether gene expression distance could also predict both types of AD on a cell subcluster level. To discover transcriptional subclusters that may provide accurate snapshots of disease, we performed gene expression-based prediction at the subcluster level. This analysis revealed Inhibitory 5 (In5) (top 3 marker genes: *CPLX3*, *TRPC3*, *KIT*) (mean AUC = 0.77 ± 0.079), Excitatory Neurons 6 (top 3 marker genes: *CA4*, *TIMP3*, *P2RY14*) (mean AUC = 0.704 ± 0.143), and Microglia 1 (top 3 marker genes: *RPL19*, *RPS11*, *RPL35*) (mean AUC = 0.669 ± 0.076) (Fig. 5a) as the most predictive cell type subclusters.

In Inhibitory Neurons 5, the top features were *RASGEF1B* and *PNISR* (Supplemental Table 9). Marker gene analysis in Excitatory 6 (Ex6) revealed a subpopulation enriched for pathways linked to Parkinson's disease, and related to translation and electron transport. We noted *LRRTM4*, which encodes leucine-rich repeat transmembrane protein 4, and *ATPB1* were top predictors in Ex6 (Supplemental Table 15). At the subcluster resolution we saw that gene expression could predict clinical disease status in 31 out of 41 subclusters (AUC > 0.5), highlighting that localized gene expression patterns may reflect AD vulnerability (Fig. 5a, Supplemental Table 8).

To further map the transcriptional axis of disease within in subclusters, we performed diffusion pseudotime analysis[38,39] by selecting a healthy root cell, based on amyloid pathology levels and cognitive diagnosis, and mapping the transcriptional progression away from this root cell (Fig. 1b). Of the 41 subclusters identified by Mathys et al [10], our pseudotime based trajectory inference stratified cells between diseased state and and healthy state Amyloid pathology in 36 subclusters (mean AUC > 0.5), including all subclusters of Inhibitory Neurons and Microglia, suggesting that gene expression within subclusters of these cells provides a predictive snapshot of Alzheimer's disease (Fig. 5b, Supplemental Table 8). To examine whether later pseudotime values actually mapped to transcriptional programs related to disease progression, we computed gene module scores for 5 Alzheimer's-related pathways, Amyloid Precursor Protein (APP) catabolism, Amyloid formation, Down regulation of Amyloid clearance, Neuroinflammatory Response, and Up regulation of Microglial activation, and correlated them with pseudotime values in each subcluster. This analysis revealed that 27 out of 41 subclusters showed a positive correlation between more disease-like pseudotime values and APP catabolism and Amyloid beta formation (Fig. 5e). 26 showed a positive correlation for negative regulation of Amyloid beta clearance and 30 showed a positive correlation for positive regulation of neuroinflammatory response (Supplemental Table 14). While 31 subclusters showed a positive correlation between pseudotime values and positive regulation of Microglia cell activation (Fig. 5e).

In Oligodendrocyte subcluster 3 (top marker genes: *HSPA1B*, *HSPA1A*, *HSP90AA1*) (Oli3), pseudotime values when used as the sole predictor of clinical AD in a logistic regression model showed a mean AUC = 0.642 (± 0.07) and an OR of 9.19 (95% CI: 8.77—9.64; FDR-adjusted q = 8.32E-44) (Fig. 5b, 5c, 5e; Supplemental Table 8). PCA analysis revealed that key genes in the pseudotime mapping of Oli3 included *PLXDC2*, *CTNND2*, *SLC5A11*, and *CDK18* (Fig. 5d). Oli3 pseudotime also classified pathology based AD well, with mean AUC = 0.66 (±0.005) and OR = 15.0 (95% CI: 13.7—16.5; FDR-adjusted q = 3.46E-63). Inhibitory Neurons 8 (top marker genes: *CHST9*, *PDGFD*, *CA1*) (In8) (mean AUC 0.62 ± 0.006; OR = 4.76, 95% CI: 4.21—5.38; FDR-adjusted q = 5.75E-33), Microglia 2 (top marker genes: *GZMB*, *PRF1*, *ITK*) (mean AUC = 0.63 ± 0.09; OR = 9.64, 95% CI: 2.78—33.42; FDR-adjusted q = 1.87E-05), and In5 (mean AUC = 0.61 ± 0.10; OR = 4.02, 95% CI: 1.25—12.85; FDR-adjusted q = 1.24E-72) all showed classification power as well. Pseudotime values in In8 (mean AUC = 0.644 ± 0.006) and In5 (mean AUC = 0.61 ± 0.10) were able to classify pathology based diagnosis as well (Fig. 5d). Across 41 studied subclusters, we found *RASGEF1B* to be a top trajectory influencing gene in 19 subclusters, with *LINGO1* and *SLC26A3* being top trajectory influencers in 16 subclusters (Fig. 5d). Additionally, there were distinct cell-type-specific genes influencing trajectories, such as *GFAP* in Astrocytes and *PLXDC2* in Oligodendrocytes (Supplementary Table 8), concordant with our predictive modeling analysis.

### Extended Figure Descriptions

**Figure 1: Overview of the TriSCOPE framework for integrative single-cell predictive and trajectory analysis in Alzheimer's disease**

a, Outline of the predictive modeling component of our TriSCOPE approach. Including cell-specific disease classification and downstream feature importance analysis.

b, Outline of the pseudotime modeling component of TriScope. We choose cells based on healthy phenotype, no Amyloid pathology (CERAD), and high cognitive diagnosis scores (COGDX). Setting the transcriptional of this healthy cell as our root, we then trace the transcriptional progression away from this cell in order to map the progression of disease. We perform this at the cell subcluster level to avoid confounding with transcriptional differences between cell types.

c, A predictive classifier trained across five splits on a global scale, no cell type differentiation, on Demographics and APOE data, shows a mean AUC of 0.528 ± 0.138. The second curve shows a predictive classifier trained across five splits on a global scale, no cell type differentiation, on Demographics, APOE, and Gene Expression data, showing a mean AUC of 0.545 ± 0.118.

d, Pyramid shows how our predictive pipeline, both in isolation and when coupled with genetic anchoring can nominate a few high-confidence gene expression targets. All found DEGs for both clinical and neuropathological Alzheimer's are listed. All predictors are also listed, followed by those GWAS-associated SNPs that overlapped with predicitively expressed genes, followed by the subset of those GWAS-associated SNPs with prefrontal tissue-specific eQTLs.

c, Heatmap showing the top 30 most predictive features from the combined classifiers trained on demographic variables (age at death, sex, education), APOE genotype, and gene expression data, separately for each major cell type: Astrocytes, Excitatory Neurons, Inhibitory Neurons, Microglia, Oligodendrocytes, and Oligodendrocyte Progenitor Cells (OPCs). Feature importance for each feature was computed as the raw importance assigned by the classifier in each of five independent splits, normalized by the total importance within each split. The square root of the mean normalized importance across splits is shown for visualization. Rows represent features (genes and demographic variables), and columns represent cell types. Color intensity indicates relative feature importance, highlighting which features most strongly contributed to Alzheimer's disease versus control classification in each cell type.
d, Circle plot summarizing a selected set of key gene features identified from SHAP analysis of combined model classifiers (Supplemental Fig. 9a) and gene expression-based classifiers, chosen for their high normalized feature importance scores and biological relevance as potential Alzheimer's disease predictors. Genes are shown for each major brain cell type: Microglia (Mic), Astrocytes (Ast), Excitatory Neurons (Exc), Inhibitory Neurons (Inh), Oligodendrocytes (Oli), and Oligodendrocyte Progenitor Cells (Opc). Nominated genes include known or emerging cell type–specific markers such as IFI44L in Microglia and GFAP in Astrocytes, as well as genes with pan–cell–type predictive value, including ARL17B, LINGO1, and RASGEF1B. This schematic highlights candidate molecular features that may underlie disease-associated transcriptional heterogeneity across cell types.
e, Bar plots showing top enriched biological pathways identified using pathway enrichment analysis of predictive genes (assigned predictive feature importance across two or more splits), performed separately for each major brain cell type: Astrocytes, Excitatory Neurons, Inhibitory Neurons, Microglia, Oligodendrocytes, and Oligodendrocyte Progenitor Cells. Enrichment was performed using g:Profiler, and odds ratios are shown on the x-axis, indicating the degree of overrepresentation of each pathway among nominated genes relative to the background gene set. Top terms highlight cell–type–specific processes potentially implicated in Alzheimer's disease–associated transcriptional changes, such as GTPase Regulator Activity and Ubiquitin Protein Ligase Binding in excitatory neurons, rho GTPase activity in microglia, and Transition Metal Ion Binding in Astrocytes.

**Figure 5: Subcluster-level predictive, trajectory, and trait enrichment analyses uncover fine-grained transcriptional programs in Alzheimer's disease**
a, Receiver operating characteristic (ROC) curves summarizing subcluster-level classification of Alzheimer's disease status using gene expression data alone. The six subclusters that exhibited the highest mean ROC performance among all subclusters analyzed. Mean area under the curve (AUC) values ± standard deviation across five cross-validation splits are indicated in the figure legends. Light blue lines represent ROC curves from individual splits, and bold lines indicate the averaged ROC curve per subcluster. Higher AUC values reflect better discrimination of Alzheimer's disease cells from control cells within each subcluster.
b, Pseudotime trajectory visualizations for eight representative subclusters with high predictive performance or biologically relevant transcriptional signatures. For each subcluster, pseudotime was inferred using diffusion pseudotime (DPT), starting from root cells selected based on advanced amyloid pathology but minimal cognitive decline. Cells are plotted in UMAP space and colored by pseudotime values (color scale: dark purple to bright yellow), representing transcriptional progression from putative early to advanced disease states. Notably, subclusters such as Oligodendrocytes 3 (Oli3), Excitatory 6 (Ex6), Inhibitory 5 (In5), and Oligodendrocytes 1 (Oli1) display strong pseudotime gradients suggestive of coherent disease-associated transcriptional trajectories. These trajectories recapitulate and extend classifier-based findings, revealing dynamic transcriptional programs aligned with AD vulnerability within each subpopulation.
c, ROC curves summarizing the predictive capacity of pseudotime trajectories for distinguishing clinical AD versus control cells across significant subclusters. Pseudotime was inferred using diffusion pseudotime (DPT) initialized from root cells with a high CERAD score and low cognitive impairment. Shown curves represent subclusters with robust mean AUC values exceeding 0.60 across seven random-seed trajectory runs. Mean AUC values ± standard error are indicated in the legend for each subcluster. Oligodendrocytes 3 (Oli3) (AUC = 0.643 ± 0.004), Inhibitory 8 (In8) (AUC = 0.630 ± 0.003), followed by Microglia 2 (Mic2), Inhibitory 5 (In5), and Inhibitory 10 (In10). The dashed line indicates chance-level classification (AUC = 0.50). These findings suggest that subcluster-specific pseudotime trajectories capture dynamic transcriptional programs strongly aligned with AD progression.
d, ROC curves summarizing the predictive capacity of pseudotime trajectories for distinguishing pathological AD versus control cells across significant subclusters. Oligodendrocytes 3 (Oli3) (AUC = 0.666 ± 0.001) once again emerged, suggesting it to be a transcriptional state that may strongly reflect both Alzheimer's pathology and clinical manifestations of the disease.
d, Smoothed expression trajectories of top trajectory-influencing genes across pseudotime in subclusters Excitatory 6, Oligodendrocytes 3, Astrocytes 0, and Inhibitory Neurons 11. For each subcluster, top genes were identified based on absolute PC4 loadings from pseudotime principal component analysis.

Expression values were smoothed using locally weighted scatterplot smoothing (LOWESS) and Z-scored for comparability. In Ex6, genes such as RASGEF1B and SLC26A3 exhibit strong monotonic trends aligned with increasing pseudotime, suggesting progressive transcriptional reprogramming associated with AD status. In Oli3, distinct dynamic trajectories were observed, including modulation of PLXDC2 expression. These profiles highlight gene-specific and subcluster-specific transcriptional programs potentially underlying differential disease vulnerability.
e, Heatmap showing Spearman correlation coefficients between pathway activity scores for five key Alzheimer's linked biological processes (APP catabolism, amyloid-β formation, negative regulation of amyloid-β clearance, positive regulation of neuroinflammatory response, and positive regulation of microglial activation) and diffusion pseudotime values across all 41 subclusters. Out of 41 subclusters, 30 subclusters showed positive correlation with neuroinflammatory response, and 31 showed positive correlation with Microglia activation.

**Supplementary Figure 1: Validation of predictive classifiers in an independent cohort**

**a,** Scatterplots comparing normalized predictive feature importance values for genes independently identified in both the Mathys et al. and Lau et al. datasets across five major brain cell types (Astrocytes, Excitatory Neurons, Inhibitory Neurons, Microglia, and Oligodendrocytes). Genes are plotted by their mean normalized importance in Mathys (x-axis) versus Lau (y-axis), with overlap counts indicated in each panel. Consistent predictive features across datasets highlight robust, cell-type–specific and pan-cell-type transcriptional determinants of Alzheimer's disease. **b,** Receiver operating characteristic (ROC) curves showing "one-shot" classification performance in the independent Lau dataset. Classifiers were first trained on the Mathys dataset using either the full feature set or incremental subsets of the top $k$ features (1–25) ranked by predictive importance in Mathys. These trained models were then frozen and directly applied to Lau without retraining. For each cell type, we report the ROC curve of the Mathys-trained model that achieved the best performance when evaluated on Lau. Results are shown for classifiers using gene expression only (blue) or gene expression plus APOE genotype (orange). **c,** Bar plots showing the top-ranked predictive features for classifiers independently trained de novo on the Lau dataset across the same five brain cell types. Normalized feature importance values highlight convergence with predictors identified in the Mathys dataset, indicating that predictive signals are stable across cohorts even when models are trained independently. **d,** Volcano plots showing differentially expressed genes (DEGs) between Alzheimer's disease and control cells across the Lau dataset, analyzed separately by cell type. Genes significant for differential expression only are shown in blue, predictive-only genes are in grey, and genes that were both predictive and differentially expressed are highlighted in red. **e,** Heatmap showing $\log_2$ fold changes of the top 80 DEGs across cell types. Genes were selected by absolute fold change and statistical significance, and ranked across cell types to visualize cross-cell-type convergence and divergence of transcriptional alterations. Color intensity indicates $\log_2$ fold change, with red = upregulation and blue = downregulation.

**Supplementary Figure 2: Differential expression analysis across combined clinical–pathological groups.**

**a,** Volcano plots showing differentially expressed genes (DEGs) across four clinical–pathological contrasts, performed separately for each of the five major brain cell types (Astrocytes, Excitatory

Neurons, Inhibitory Neurons, Microglia, Oligodendrocytes, and Oligodendrocyte Progenitor Cells). The four contrasts tested were: (i) Clinical AD versus Clinical Control within pathology-negative individuals, (ii) Pathology AD versus Pathology Control within clinically non-demented individuals, (iii) Clinical AD versus Clinical Control within pathology-positive individuals, and (iv) Pathology AD versus Pathology Control within clinically diagnosed AD cases. Genes significant for differential expression (FDR-adjusted $P < 0.05$) are colored in red (upregulated) and blue (downregulated). **b,** Pathway enrichment analysis of DEGs identified in Microglia for the contrast of Pathology-positive Clinical AD versus Pathology-positive Clinical Controls. Top enriched biological processes included regulation of neuroinflammatory response, positive regulation of cytokine-mediated signaling, and microglial activation pathways. **c,** Pathway enrichment analysis of DEGs identified in Oligodendrocytes for the contrast of Pathology-positive Clinical AD versus Pathology-positive Clinical Controls. Enriched pathways included cellular responses to unfolded protein, regulation of myelination, and autophagy-related processes, highlighting potential oligodendrocyte-specific mechanisms of disease progression.

**Supplementary Figure 3: Demographic, compositional, and clustering characteristics of Mathys and Lau datasets.**

**a,** Distribution of donors across clinical and neuropathological categories in the Mathys dataset, showing separation into four combined groups (Clinical Control/Pathology Control, Clinical AD/Pathology Control, Clinical Control/Pathology AD, and Clinical AD/Pathology AD). **b,** Cell type composition across both Mathys and Lau datasets, demonstrating broadly consistent representation of astrocytes, excitatory neurons, inhibitory neurons, microglia, and oligodendrocytes. **c,** Age-at-death distributions for AD cases and controls across both cohorts, indicating similar demographic ranges. **d,** UMAP projection of the validation (Lau) dataset annotated using marker genes from the original study, showing robust separation of major brain cell types. **e,** Validation dataset annotated with Mathys marker genes, demonstrating concordance in broad cell type assignment with panel D. **f,** Leiden clustering of the Lau dataset, capturing additional substructure within annotated cell types. **g,** UMAP projection of the Mathys dataset annotated with broad cell types. **h,** Subcluster-level annotation of the Mathys dataset, showing finer stratification within major cell classes. **i,** Leiden clustering of the Mathys dataset, highlighting resolution of transcriptionally distinct subpopulations. **j,** Post-mortem interval (PMI) distributions for clinical AD versus control donors in the Mathys dataset, showing no significant difference between groups. **k,** PMI distributions for clinical AD versus control donors in the Lau dataset, similarly showing no significant difference.

Supplementary Figure 9: SHAP-based interpretation of predictive models for clinical AD status stratified by APOE genotype

**a**, Bar plots showing the top gene-level predictors of clinical AD classification across major brain cell types identified by SHAP analysis. Genes are ranked by normalized SHAP importance, with representative high-confidence features such as RASGEF1B (excitatory neurons), HSPH1 (excitatory neurons), and IFI44L (microglia). These features informed the set of genes highlighted in the two-dimensional SHAP plots in panel B. **b**, Heatmaps displaying per-gene SHAP ranks across APOE genotype groups (non-carriers, heterozygous ε4, homozygous ε4/ε4), illustrating consistency and divergence of predictive features under distinct genetic risk backgrounds. **c**, Radar plots summarizing APOE-stratified SHAP values for selected top genes within each major cell type, showing shifts in predictive contribution between genotype-defined subgroups. **d**, Heatmap showing Spearman correlation between gene expression and predicted AD probability across cell types for a representative subset of key predictors (e.g., RASGEF1B, LINGO1, ARL17B, MT2A, APOC1). Positive correlations highlight cell-type–specific associations between expression level and model-predicted AD probability, whereas negative correlations suggest context-dependent or compensatory roles.

Supplementary Figure 10: Linking cell-type–specific predictive features to AD GWAS and brain eQTL support.

**a,** Heatmap of clinical AD predictors (identified in our modeling framework) that overlap with genome-wide significant AD GWAS loci ($P < 5 \times 10^{-8}$) and additionally exhibit support from brain tissue eQTL datasets. Predictors are shown in a cell-type–specific manner, though the eQTL evidence derives from bulk brain tissue rather than cell-type–specific eQTL resources. **b,** Heatmap of AD neuropathology predictors overlapping with GWAS loci and brain eQTL support, analogous to panel A. **c,** Heatmaps showing $-\log_{10}$ GWAS P-values for top neuropathology-associated predictors overlapping with GWAS loci, stratified by cell type. Up to 10 most significant genes are shown per cell type. Notably, APOE exhibits strong GWAS association in both astrocytes and microglia. **d,** Venn diagrams illustrating the overlap between clinical AD GWAS-supported predictors and neuropathology GWAS-supported predictors across cell types, highlighting convergent and distinct sets of genetically anchored disease-relevant features.
